# Effectiveness of health mediation to promote organized cancer screening among underserved and under-screened populations in Marseille, France: findings from a repeated cross-sectional survey

**DOI:** 10.64898/2026.03.06.26347781

**Authors:** Eva Legendre, Anne Dutrey-Kaiser, Yazid Attalah, Ghislaine Boyer, Steve Nauleau, Jean Gaudart, David Kelly, Céline Caserio-Schönemann, Philippe Malfait, Pascal Chaud, Lauriane Ramalli, Chrystelle Gastaldi, Florian Franke, Stanislas Rebaudet

## Abstract

**Background:** Although health mediation is widely studied in the U.S. through community health worker programs, evidence on their effectiveness in promoting cancer screening in Europe is limited. Since 2022, the “*13 en Santé*” program has implemented a multicomponent health mediation intervention — combining educational activities, outreach strategies, and navigation support — in socioeconomically disadvantaged neighbourhoods of Marseille, France. This study evaluates the effectiveness of this program in promoting breast, colorectal, and cervical cancer screening.

**Methods:** A controlled before-after design based on two cross-sectional surveys was conducted in 2022 and 2024 in intervention or control neighbourhoods. Individuals aged 18–74 were randomly selected and interviewed via door-to-door questionnaires. Weighting was applied to account for stratified sampling and to align age and sex distributions with census data. Weighted logistic regression models were fitted for each cancer screening to estimate the intervention’s effects on uptake and awareness at both individual and population levels.

**Findings:** Overall, 4,523 individuals were included across the two cross-sectional surveys. The program successfully reached individuals facing cumulative socioeconomic barriers to healthcare access. No significant population-level effect was observed. At the individual level, declared exposure to health mediation was associated with significantly higher uptakes of breast and colorectal cancer screenings (breast: 54% vs. 74%, OR=2.3 [1.1–4.5]; colorectal: 30% vs. 50%, OR=2.8 [1.3–5.8]). In addition, colorectal cancer screening awareness was significantly higher among exposed participants (83% vs. 93%, OR=8.1 [2.1–31]).

**Interpretation:** This study provides the first evidence that a multicomponent health mediation intervention could effectively promote breast and colorectal cancer screening in disadvantaged French neighbourhoods. The study highlights screening-specific mechanisms of action that should be considered to further optimize intervention effectiveness.

**Funding:** The survey was funded by the Regional Health Agency of Provence–Alpes–Côte d’Azur and Santé publique France.

## Introduction

Reducing social inequalities in health is essential to progress through a more equitable and just society. However, these disparities have increased over the past decade in Europe, particularly in France^1,2^. Community health workers (CHWs), predominantly implemented and evaluated in low- and middle-income countries and in high-income Anglo-American settings, are increasingly recognized in Europe as a promising intervention to mitigate social health inequalities.^3^

To address its significant territorial and social inequities, France has defined ‘priority neighbourhoods’ (*quartiers prioritaires de la politique de la ville*) focusing resources on areas with high socioeconomic disadvantage^4^. In Marseille, the country’s second-largest city, 28% of the population (243,471 individuals) reside in a priority neighbourhood^5^. These neighbourhoods are concentrated in the city centre (i.e., 1^st^ to 3^rd^ arrondissements) and the northern arrondissements (i.e., 13^th^ to 16^th^ arrondissements), creating a North–South divide. Residents in these priority neighbourhoods have less access to healthcare (e.g., primary care physician, universal health insurance coverage) and preventive services (e.g., cancer screening, vaccination), compared with the rest of the city^6^.

Additionally, to reduce health inequalities in the central and northern arrondissements of Marseille, the health mediation program called “*13 en Santé*” was launched in priority neighbourhoods in October 2022 and is still ongoing in 2026. In France, health authorities conceptualise health mediation as an expansion of the community health worker (CHW) approach, focusing not on the actors themselves but on the goals and functions of the intervention. Health mediation acts as a bridge between underserved populations and the healthcare system, to improve access to health rights, prevention, care and is delivered by health mediators, who may include CHWs^7^. The “*13 en Santé*” program offers culturally appropriate education sessions or information through outreach interventions, as well as social and health navigation tailored to individual’s autonomy and needs. Outreach strategies include door-to-door visits, street stands, or workshops conducted with partner organizations — either outside or within healthcare facilities — to reach a broad population, particularly those most distant from the healthcare system. The program aims to promote access to healthcare and preventive services, including population-based cancer screenings programs.

In France, cancer is the leading cause of death among men and the second leading cause among women^8^. France offers free screening programs for breast cancer (mammography targeting women aged 50–74 years, every two years), colorectal cancer (faecal immunochemical test for adults aged 50–74 years, every two years), and uterine cervical cancer (two Pap smears one-year apart and a third three years later for women aged 25–29 years, then a high risk human papillomavirus (HR-HPV) PCR every five years for women aged 30-64)^9^. Eligible individuals receive an invitation letter by mail to participate in cancer screening.

Despite the availability of these cancer screening programs, free of charge for all individual with universal health coverage, uptake has shown substantial territorial and social disparities, as seen in Marseille. First, screening rates in Marseille are consistently lower than regional and national levels; for example, most recent colorectal cancer screening uptake rates reached 25.1% in Marseille, compared with 30.9% regionally and 34.2% nationally^10,11^. Second, differences were observed across screening programs at the city level, with uptake ranging from 25.4% for colorectal cancer (2021–2022) to 47.5% for cervical cancer (2020–2022) and 58.2% for breast cancer (2021–2022)^10^. Third, disparities following clear geographic and socioeconomic gradients were also observed within the city. In 2021–2022, breast cancer screening uptake in the central and northern arrondissements was estimated at 52.8% among individuals with paid complementary health insurance, and 39% among those with subsidized complementary health insurance that is provided to people with the lowest incomes^10,12^. Conversely, in the southern part of the city, uptakes reached 64.2% and 44%, respectively^10^. Accordingly, and in alignment with regional health authorities, the promotion of population-based cancer screening programs was established as a priority of the “*13 en Santé*” health mediation program.

The effectiveness of CHW programs—or of navigation, educational, or outreach interventions that constitute core components of CHW models—in promoting cancer screening uptake has been predominantly investigated in the United States, particularly through randomized controlled trials^13,14^. But CHW interventions generally targeted specific ethnic communities or a single cancer screening program. Moreover, their efficacy may be influenced by contextual factors, such as the healthcare system organization or cancer screening program^13,14^. In the United Kingdom, a community health and wellbeing worker intervention promoting vaccination, cancer screening, and access to National Health Service services was implemented at the household level in a deprived inner-London borough in 2021–2022. Preliminary findings indicated a 40% higher service uptake in the intervention group compared with the control group after six months of activity^15^.

In continental Europe, evidence has remained scarce. To date, only one study conducted in France tested a patient navigation intervention targeting the general population to improve colorectal cancer screening uptake in 2011-2013^16^. The study showed that, when implemented across all social strata, it modestly increased overall participation (OR=1.19 [95% CI: 1.10–1.29], 3.2% increase in participation rate), but also exacerbated social inequalities in screening rates^16^.

This study aims to evaluate the effectiveness of a health mediation program —combining educational, outreach, and navigation strategies — in promoting breast, colorectal, and cervical cancer screening among underserved and under-screened populations living in socioeconomically disadvantaged neighbourhoods in Marseille, France, representing, to our knowledge, the first such study in Europe.

## Methods

### Setting

At the request of the regional health authority (*Agence Régionale de Santé Provence–Alpes–Côte d’Azur, ARS-Paca*), the “*13 en Santé*” health mediation program has been implemented since October 2022 in Marseille by two operators: CORHESAN, supported by a private nonprofit hospital, Hôpital Européen, and a non-profit organization, Prospective & Coopération, targeting nine priority neighbourhoods of the city centre; and SEPT, a non-profit organization, targeting four priority neighbourhoods of the northern arrondissements^17^. Teams were composed of trained health mediators and nurses. They promote breast, cervical and colorectal cancer screenings through educational sessions with various outreach strategies, tailored information using motivational interviewing, and navigation support to mammography centres, gynaecological consultations, or distribution of faecal occult blood testing kits (**Sup. method 1 p.5**).

### Study design

In parallel to this health mediation program, health authorities also commanded an assessment of the intervention effectiveness primarily on cancer screening uptake and secondarily on participants’ awareness. This was conducted using a before-and-after, control-intervention design based on two repeated cross-sectional surveys conducted in 13 intervention and 26 control neighbourhoods. Control neighbourhoods were defined as areas without planned health mediation activities and were selected to be socioeconomically like the intervention neighbourhoods (**Sup. method 2**, **Sup. figure 1 p.5-6**). The baseline survey (T0) was conducted before the start of the health mediation program from 18 August to 30 September 2022 in intervention neighbourhoods, and from 28 August to 27 December 2022 in control areas. The follow-up survey (T1) was conducted one year and half after the start of the health mediation program in two phases due to operational reasons: the first phase (T1 P1) from 22 April to 20 June 2024, and the second phase (T1 P2) from 9 to 24 September 2024.

The anonymized questionnaires were validated by the Data protection officer of Santé publique France (SpF) prior to the survey in accordance with the European General Data Protection Regulation. The study was approved by the Hôpital Européen ethics committee (No. 2025-08-05-13S).

### Participants

Residents aged 18 to 74 years in both intervention or control neighbourhood were randomly selected through door-to-door visits (**Sup. method 3 p.6**). While only individuals aged 25 to 74 years were eligible for at least one of the targeted cancer screening programs, the 18–24-year-old group was also included to evaluate vaccination outcomes. This article focuses on results related to cancer screening promotion.

### Procedures

A knowledge-attitude-practice questionnaire on cancer screening and vaccination was administered to participants. Outcomes related to breast, cervical, and colorectal cancer screening awareness and uptake (i.e. screening up to date according to national guidelines) were collected from participants using tablets. Individual information regarding socioeconomic characteristics (i.e., self-reported gender, age, birthplace, mother tongue, education level, occupational status, residential status) and general access to healthcare system (i.e., primary care physician, receipt of cancer screening invitation) was also collected. During the T1 follow-up survey, participants were also asked whether they had encountered a CORHESAN or SEPT health mediator in the past and whether cancer screening programs and vaccination had been discussed (**Sup. table 1 p.7-8**).

### Statistical analysis

Statistical analyses were conducted using R 4.2.2 (R Development Core Team, R Foundation for Statistical Computing, Vienna, Austria) and {ade4}, {survey}, {lme4} and {rpart} packages^18–20^.

#### Study size

The sample size at baseline (T0) and follow-up (T1) was calculated using the estimated baseline cancer screening uptake rates in the targeted arrondissements (46% for breast, 18% for colorectal, and 32% for uterine cervical cancer)^10,21^. An expected absolute increase of 10% in uptake was assumed. Calculations accounted for the population structure of both intervention and control neighbourhoods, including overlap among eligible groups^22^. Sample size estimation was performed for a two-sided test with a type I error rate of 5% and a statistical power of 80%. Participant recruitment was stratified by age, gender, and neighbourhood.

#### Socioeconomic profiles construction

To capture cumulative socioeconomic advantages or disadvantages, we constructed participants profiles that summarized key characteristics (birthplace, mother tongue, education level, occupational status, residential status). A socioeconomic profile was defined as a cluster of individuals sharing similar socioeconomic patterns. To create these profiles, a multiple component analysis followed by a hierarchical ascendant clustering was performed. Profile imputation was carried out for individuals with at least one available socioeconomic characteristic using a decision tree built through recursive partitioning, which enabled assigning individuals to the most probable profile based on their available characteristics.

#### Weighting based on census data and stratification sampling

Individual weighting was applied to account for the three-level stratification and the distribution of the general population by age and sex in the included neighbourhoods, based on census data and using {survey} package^22^. All percentages were estimated using the applied survey weights.

#### Weighted modelling

Weighted logistic regression models were performed separately for breast, colorectal, and cervical cancer screenings to estimate the effect of the health mediation intervention on screening uptake and awareness at both the population and individual levels. Population-level effects were estimated by comparing screening uptake or knowledge between control and intervention neighbourhoods, before and after the implementation of the intervention. Individual-level effects were estimated by comparing participants who reported having encountered a health mediator with those who had not, regardless of their home neighbourhood. Effect estimates were adjusted for individual confounding factors in multivariate models, including age, socioeconomic profile, access to a primary care physician, receipt of the screening invitation letter, residential arrondissement, and gender, when appropriate.

Secondary weighted multivariate analyses were conducted to highlight individual characteristics associated with general access to healthcare (i.e., primary care physician, receipt of the screening invitation letter) and with reporting an encounter with a health mediator.

## Results

### Participants description

Overall, 4,530 individuals participated in both cross-sectional surveys. Seven questionnaires were excluded due to missing neighbourhood information. Thus, 4,523 individuals were included in the final analysis of the two cross-sectional surveys and 4,095 (90.54%) were eligible to at least one cancer screening (**Table 1**). Four socioeconomic profiles were identified and labelled based on their predominant characteristics: “Native speaker renter” (NS renter) regrouping 42.85% of the participants (n=1938); “Non-native speaker renter” (non-NS renter), with 35.55% (n=1608); “Native speaker homeowner” (NS homeowner) accounting for 10.90% (n=493); and “Native speaker student” (NS student), representing 10.55% (n=477) (**Table 1**, **Sup. Figures 2-7 p.9-14**).

**Table 1.** Baseline and follow-up characteristics of participants in control and intervention neighbourhoods (unweighted), n (%).

|  | Baseline (T0) |  | Follow-up (T1) |  | Overall<br>(n=4523) |
| --- | --- | --- | --- | --- | --- |
|  | Control<br>(n=1384,<br>30.60%) | Intervention<br>(n=1261,<br>27.88%) | Control<br>(n=981,<br>21.69%) | Intervention<br>(n=897,<br>19.83%) |  |
| **Operational information** |  |  |  |  |  |
| **Survey phases** |  |  |  |  |  |
| P0 (August-Dec. 2022) | 1384 (100.00) | 1261 (100.00) | 0 (0.00) | 0 (0.00) | 2645 (58.48) |
| P1 (April-June 2024) | 0 (0.00) | 0 (0.00) | 889 (90.62) | 583 (64.99) | 1472 (32.54) |
| P2 (Sept. 2024) | 0 (0.00) | 0 (0.00) | 81 (8.26) | 281 (31.33) | 362 (8.00) |
| Missing data | 0 (0.00) | 0 (0.00) | 11 (1.12) | 33 (3.68) | 44 (0.97) |
| **Demographic characteristics** |  |  |  |  |  |
| **Gender** |  |  |  |  |  |
| Man | 544 (39.31) | 500 (39.65) | 325 (33.13) | 317 (35.34) | 1686 (37.28) |
| Woman | 840 (60.69) | 761 (60.35) | 655 (66.77) | 579 (64.55) | 2835 (62.68) |
| Missing data<sup>1</sup> | 0 (0.00) | 0 (0.00) | 1 (0.10) | 1 (0.11) | 2 (0.04) |
| **Age** |  |  |  |  |  |
| 18-24 | 139 (10.04) | 138 (10.94) | 81 (8.26) | 70 (7.80) | 428 (9.46) |
| 25-29 | 146 (10.55) | 107 (8.49) | 99 (10.09) | 87 (9.70) | 439 (9.71) |
| 30-34 | 108 (7.80) | 113 (8.96) | 69 (7.03) | 63 (7.02) | 353 (7.80) |
| 35-39 | 130 (9.39) | 129 (10.23) | 68 (6.93) | 64 (7.13) | 391 (8.64) |
| 40-44 | 92 (6.65) | 106 (8.41) | 53 (5.40) | 50 (5.57) | 301 (6.65) |
| 45-49 | 112 (8.09) | 95 (7.53) | 61 (6.22) | 59 (6.58) | 327 (7.23) |
| 50-54 | 153 (11.05) | 158 (12.53) | 143 (14.58) | 143 (15.94) | 597 (13.20) |
| 55-59 | 141 (10.19) | 117 (9.28) | 98 (9.99) | 83 (9.25) | 439 (9.71) |
| 60-64 | 110 (7.95) | 96 (7.61) | 117 (11.93) | 110 (12.26) | 433 (9.57) |
| 65-69 | 106 (7.66) | 88 (6.98) | 90 (9.17) | 89 (9.92) | 373 (8.25) |
| 70-74 | 147 (10.62) | 114 (9.04) | 102 (10.40) | 79 (8.81) | 442 (9.77) |
| **Arrondissement** |  |  |  |  |  |
| 1<sup>st</sup> | 86 (6.21) | 206 (16.34) | 69 (7.03) | 170 (18.95) | 531 (11.74) |
| 2<sup>nd</sup> | 132 (9.54) | 183 (14.51) | 91 (9.28) | 138 (15.38) | 544 (12.03) |
| 3<sup>rd</sup> | 461 (33.31) | 240 (19.03) | 339 (34.56) | 157 (17.50) | 1197 (26.46) |
| 13<sup>th</sup> | 208 (15.03) | 185 (14.67) | 155 (15.80) | 148 (16.50) | 696 (15.39) |
| 14<sup>th</sup> | 111 (8.02) | 131 (10.39) | 64 (6.52) | 89 (9.92) | 395 (8.73) |
| 15<sup>th</sup> | 243 (17.56) | 162 (12.85) | 156 (15.90) | 105 (11.71) | 666 (14.72) |
| 16<sup>th</sup> | 143 (10.33) | 154 (12.21) | 107 (10.91) | 90 (10.03) | 494 (10.92) |
| **Socio-economic characteristics** |  |  |  |  |  |
| **Birthplace** |  |  |  |  |  |
| France | 872 (63.01) | 633 (50.20) | 574 (58.51) | 500 (55.74) | 2579 (57.02) |
| Abroad | 506 (36.56) | 621 (49.25) | 398 (40.57) | 395 (44.04) | 1920 (42.45) |
| Missing data | 6 (0.43) | 7 (0.56) | 9 (0.92) | 2 (0.22) | 24 (0.53) |
| **Mother tongue** |  |  |  |  |  |
| French | 868 (62.72) | 643 (50.99) | 641 (65.34) | 572 (63.77) | 2724 (60.23) |
| Other | 506 (36.56) | 608 (48.22) | 331 (33.74) | 323 (36.01) | 1768 (39.09) |
| Missing data | 10 (0.72) | 10 (0.79) | 9 (0.92) | 2 (0.22) | 31 (0.69) |
| **Education level** |  |  |  |  |  |
| Primary school | 171 (12.36) | 198 (15.70) | 57 (5.81) | 105 (11.71) | 531 (11.74) |
| Secondary school | 685 (49.49) | 564 (44.73) | 407 (41.49) | 328 (36.57) | 1984 (43.86) |
| Undergraduate | 382 (27.60) | 371 (29.42) | 384 (39.14) | 336 (37.46) | 1473 (32.57) |
| Postgraduate | 102 (7.37) | 76 (6.03) | 95 (9.68) | 68 (7.58) | 341 (7.54) |
| Other | 26 (1.88) | 29 (2.30) | 22 (2.24) | 46 (5.13) | 123 (2.72) |
| Missing data | 18 (1.30) | 23 (1.82) | 16 (1.63) | 14 (1.56) | 71 (1.57) |
| **Occupational status** |  |  |  |  |  |
| Employed | 566 (40.90) | 468 (37.11) | 400 (40.77) | 370 (41.25) | 1804 (39.89) |
| Student | 89 (6.43) | 82 (6.50) | 62 (6.32) | 54 (6.02) | 287 (6.35) |
| Homemaker | 156 (11.27) | 242 (19.19) | 154 (15.70) | 141 (15.72) | 693 (15.32) |
| Retired | 292 (21.10) | 236 (18.72) | 216 (22.02) | 209 (23.30) | 953 (21.07) |
| Unemployed | 154 (11.13) | 147 (11.66) | 70 (7.14) | 71 (7.92) | 442 (9.77) |
| Other | 87 (6.29) | 62 (4.92) | 56 (5.71) | 45 (5.02) | 250 (5.53) |
| Missing data | 40 (2.89) | 24 (1.90) | 23 (2.34) | 7 (0.78) | 94 (2.08) |
| **Residential status** |  |  |  |  |  |
| Homeowner | 192 (13.87) | 122 (9.67) | 182 (18.55) | 104 (11.59) | 600 (13.27) |
| Renter | 989 (71.46) | 936 (74.23) | 684 (69.72) | 653 (72.80) | 3262 (72.12) |
| Family | 142 (10.26) | 143 (11.34) | 86 (8.77) | 114 (12.71) | 485 (10.72) |
| Other | 47 (3.40) | 33 (2.62) | 16 (1.63) | 22 (2.45) | 118 (2.61) |
| Missing data | 14 (1.01) | 27 (2.14) | 13 (1.33) | 4 (0.45) | 58 (1.28) |
| **Socioeconomic profile<sup>2</sup>** |  |  |  |  |  |
| Native speaker homeowner | 159 (11.49) | 88 (6.98) | 156 (15.90) | 90 (10.03) | 493 (10.90) |
| Native speaker renter | 642 (46.39) | 483 (38.30) | 426 (43.43) | 387 (43.14) | 1938 (42.85) |
| Native speaker student | 150 (10.84) | 139 (11.02) | 86 (8.77) | 102 (11.37) | 477 (10.55) |
| Non-native speaker renter | 432 (31.21) | 550 (43.62) | 309 (31.50) | 317 (35.34) | 1608 (35.55) |
| Missing data | 1 (0.07) | 1 (0.08) | 4 (0.41) | 1 (0.11) | 7 (0.15) |
| **Health mediation** |  |  |  |  |  |
| **Encounter with a health mediator** |  |  |  |  |  |
| Not reported | 1384 (100.00) | 1261 (100.00) | 896 (91.34) | 748 (83.39) | 4289 (94.83) |
| Reported | 0 (0.00) | 0 (0.00) | 46 (4.69) | 117 (13.04) | 163 (3.60) |
| Missing data | 0 (0.00) | 0 (0.00) | 39 (3.98) | 32 (3.57) | 71 (1.57) |
| **Access to healthcare system** |  |  |  |  |  |
| **Primary care physician** |  |  |  |  |  |
| Without | 196 (14.16) | 175 (13.88) | 83 (8.46) | 55 (6.13) | 509 (11.25) |
| With | 1183 (85.48) | 1085 (86.04) | 894 (91.13) | 840 (93.65) | 4002 (88.48) |
| Missing data | 5 (0.36) | 1 (0.08) | 4 (0.41) | 2 (0.22) | 12 (0.27) |
| **Breast cancer screening invitation (women aged 50–74 years, n=1438)** |  |  |  |  |  |
| Not received | 45 (11.42) | 49 (14.63) | 56 (14.74) | 44 (13.37) | 194 (13.49) |
| Received | 318 (80.71) | 248 (74.03) | 307 (80.79) | 267 (81.16) | 1140 (79.28) |
| Missing data | 31 (7.87) | 38 (11.34) | 17 (4.47) | 18 (5.47) | 104 (7.23) |
| **Colorectal cancer screening invitation (men and women aged 50–74 years, n=2284)** |  |  |  |  |  |
| Not received | 117 (17.81) | 137 (23.91) | 145 (26.36) | 147 (29.17) | 546 (23.91) |
| Received | 470 (71.54) | 363 (63.35) | 355 (64.55) | 300 (59.52) | 1488 (65.15) |
| Missing data | 70 (10.65) | 73 (12.74) | 50 (9.09) | 57 (11.31) | 250 (10.95) |
| **Cervical cancer screening invitation (women aged 25–64 years, n=2118)** |  |  |  |  |  |
| Not received | 187 (30.11) | 225 (38.93) | 158 (32.92) | 117 (26.65) | 687 (32.44) |
| Received | 300 (48.31) | 239 (41.35) | 278 (57.92) | 271 (61.73) | 1088 (51.37) |
| Missing data | 134 (21.58) | 114 (19.72) | 44 (9.17) | 51 (11.62) | 343 (16.19) |
| **Cancer screening uptake** |  |  |  |  |  |
| **Breast cancer (women aged 50–74 years, n=1438)** |  |  |  |  |  |
| Up to date | 191 (48.48) | 181 (54.03) | 224 (58.95) | 142 (43.16) | 738 (51.32) |
| Not up to date | 188 (47.72) | 140 (41.79) | 147 (38.68) | 176 (53.50) | 651 (45.27) |
| Missing data | 15 (3.81) | 14 (4.18) | 9 (2.37) | 11 (3.34) | 49 (3.41) |
| **Colorectal cancer (men and women aged 50–74 years, n=2284)** |  |  |  |  |  |
| Up to date | 209 (31.81) | 170 (29.67) | 170 (30.91) | 129 (25.60) | 678 (29.68) |
| Not up to date | 423 (64.38) | 375 (65.45) | 363 (66.00) | 367 (72.82) | 1528 (66.90) |
| Missing data | 25 (3.81) | 28 (4.89) | 17 (3.09) | 8 (1.59) | 78 (3.42) |
| **Cervical cancer (women aged 25–64 years, n=2118)** |  |  |  |  |  |
| Up to date | 404 (65.06) | 394 (68.17) | 340 (70.83) | 270 (61.50) | 1408 (66.48) |
| Not up to date | 182 (29.31) | 144 (24.91) | 125 (26.04) | 142 (32.35) | 593 (28.00) |
| Missing data | 35 (5.64) | 40 (6.92) | 15 (3.12) | 27 (6.15) | 117 (5.52) |
| **Cancer screening awareness** |  |  |  |  |  |
| **Breast cancer (women aged 50–74 years, n=1438)** |  |  |  |  |  |
| Aware | 379 (96.19) | 312 (93.13) | 370 (97.37) | 307 (93.31) | 1368 (95.13) |
| Not aware | 13 (3.30) | 23 (6.87) | 7 (1.84) | 21 (6.38) | 64 (4.45) |
| Missing data | 2 (0.51) | 0 (0.00) | 3 (0.79) | 1 (0.30) | 6 (0.42) |
| **Colorectal cancer** (men and women aged 50–74 years, n=2284) |  |  |  |  |  |
| Aware | 562 (85.54) | 456 (79.58) | 481 (87.45) | 386 (76.59) | 1885 (82.53) |
| Not aware | 93 (14.16) | 116 (20.24) | 63 (11.45) | 116 (23.02) | 388 (16.99) |
| Missing data | 2 (0.30) | 1 (0.17) | 6 (1.09) | 2 (0.40) | 11 (0.48) |
| **Cervical cancer** (women aged 25–64 years, n=2118) |  |  |  |  |  |
| Aware | 524 (84.38) | 472 (81.66) | 427 (88.96) | 386 (87.93) | 1809 (85.41) |
| Not aware | 94 (15.14) | 105 (18.17) | 46 (9.58) | 48 (10.93) | 293 (13.83) |
| Missing data | 3 (0.48) | 1 (0.17) | 7 (1.46) | 5 (1.14) | 16 (0.76) |

<sup>1</sup>, missing data corresponded to a participant declaring undifferentiated gender, but due to low total headcount and statistical limit, we couldn't consider this category in our analysis.
<sup>2</sup>, variable constructed from the following socioeconomic characteristics: birthplace, mother tongue, educational level, occupational status and residential status.

### Weighted sample

Women aged over 40 years were overrepresented compared with the general population (**Sup. figure 8 p.15**). To correct for sampling imbalance, a weighting was applied to each participant —assigning higher weights to younger individuals and to men — which restored an age and sex distribution comparable to the underlying population in each neighbourhood (**Sup. figures 9-10 p.16-17**). Weighting slightly modified the distribution of socioeconomic profiles, increasing the proportion of NS students in the T1 neighbourhoods. (**Sup. figure 11 p.18**).

### Access to primary care physician and reception of screening invitation letters

Most participants (weighted percentage (WP)=85.14%, n=4,000) reported having access to a primary care physician (**Sup. table 2 p.19**). Access to a physician was significantly lower among non-NS renters (WPs=82.07%, n=1403) compared with NS renters (WPs=88.81%, n=1764, OR=0.58 [0.42-0.79]) (**Sup. table 2 p.19, sup. figure 12 p.20**).

Overall, reported invitation letter receipt was highest for breast cancer screening, with 80.97% of women (n=1,140), followed by colorectal cancer screening at 65.15% (n=1,488), and cervical cancer screening (WP=53.30%, n=1,088) (**Sup. tables 3-5 P.21,23,25**). For all three cancer screenings, non-NS renters consistently reported significantly lower invitation letter receipt compared with NS renters (breast: WPs=77.31% vs 89.55%, OR=0.38 [0.24-0.60]; colorectal: WPs=57.75% vs 77.78%, OR=0.39 [0.28-0.55]; cervical: WPs=52.41% vs 69.33%, OR=0.45 [0.33-0.61]) (**Sup. tables 3-5, sup. figures 13-15 P.21-26**). On the contrary, individuals who reported access to a primary care physician significantly reported higher invitation letter receipt compared with those who did not reported access to a physician (breast: WPs=86.68% vs 63.50%, OR=4.46 [2.24-8.89]; colorectal: WPs=73.73% vs 48.36%, OR=3.29 [1.85-5.85]; cervical: WPs=63.63% vs 40.21%, OR=2.57 [1.59-4.14]). Additionally, encountering a health mediator was significantly associated with receipt of the screening invitation only for cervical cancer (WP=78.80%, n=65), compared with participants who did not report such an encounter (WP=60.93%, n=1,008, OR=2.00 [1.03-3.88]).

### Health mediator encounter

In the T1 survey, 12.78% (n=117) of participants in the intervention neighbourhoods reported having encountered a health mediator, with exposure ranging from 18.21% (n=92) in the central arrondissements to 5.53% (n=25) in the northern neighbourhoods. In the control neighbourhoods, the encounter rate was 3.66% (n=46). Reported health mediator encounters were significantly more frequent in the T1 P1 intervention neighbourhoods (WP=16.62%, n=96) than in the T1 P1 control neighbourhoods (WP=3.80%, n=42, OR=5.43 [3.10-9.51]) (**Figure 1, Sup. table 6 p.27**). In contrast, no significant difference was observed between the T1 P2 intervention neighbourhoods (WP=3.65%, n=14) and T1 controls (OR=0.96 [0.43-2.12]). Non-NS renters (WP=10.81%, n=88, OR=2.84 [1.63-4.96]) and NS students (WP=6.26%, n=15, OR=2.80 [1.25-6.27]) were significantly more likely to report health mediator encounter than NS renters (WP=3.68%, n=44).

**Figure 1.**
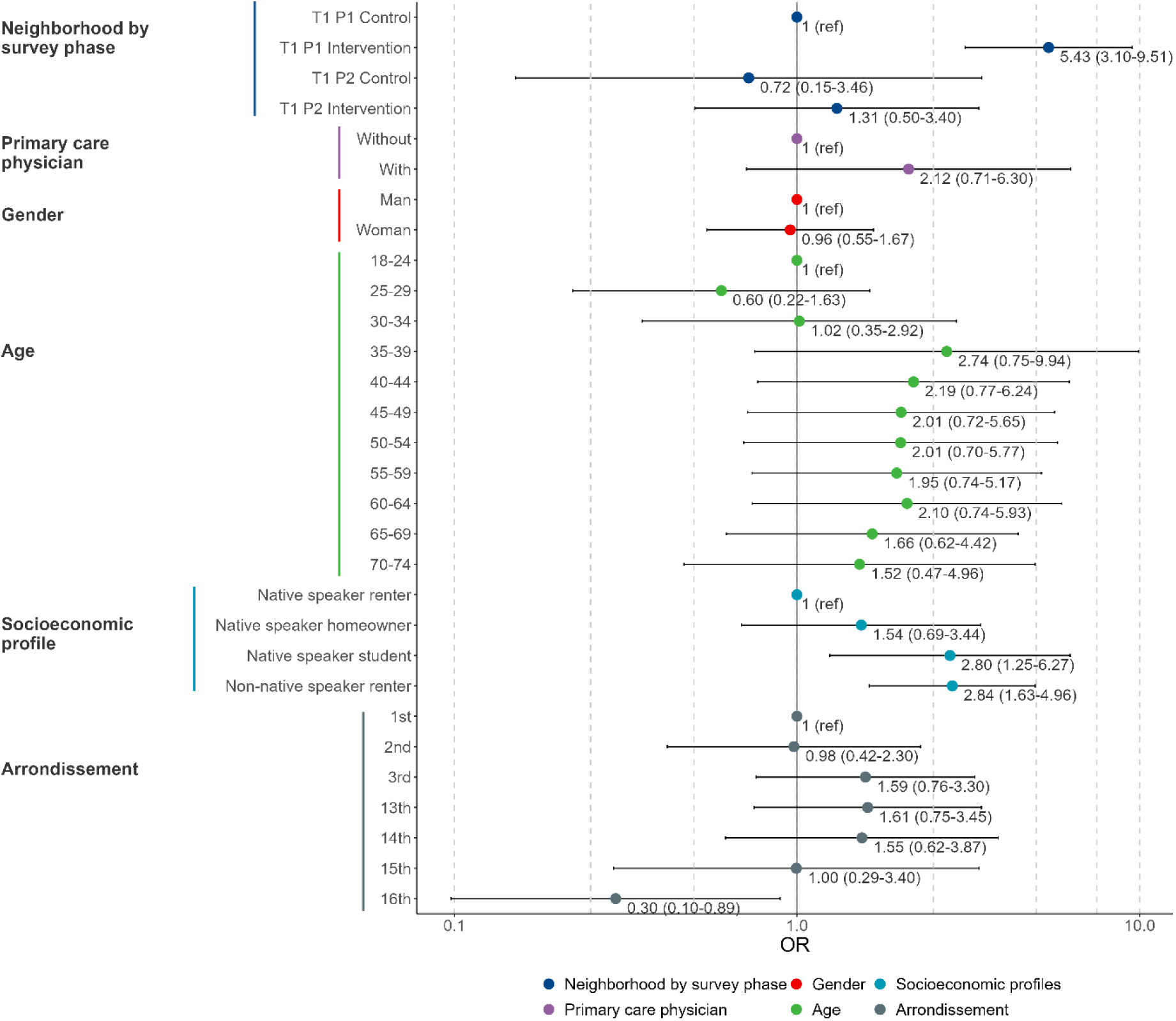
Factors associated with participants reporting an encounter with a health mediator during the T1 survey, in the weighted multivariate analysis.

### Population-level effect of health mediation on cancer screening uptake

No effect of the health mediation program on cancer screening uptake was observed when exposure was defined at the neighbourhood level: for all three cancers, uptake rates remained stable over time — from T0 to T1— and did not differ between intervention and control neighbourhoods (**Figure 2A**, **Figure 3, Sup. tables 7-9 p.28-30**). Overall, reported uptake was highest for cervical cancer screening (WP=71.34%, n=1408), followed by breast cancer screening (WP=54.48%, n=738), and substantially lower for colorectal cancer screening (WP=29.90%, n=678). No population-effect of health mediation was also observed on cancer screening awareness (**Sup. tables 10-12, sup. figures 16-18 p.31-36**).

**Figure 2.**
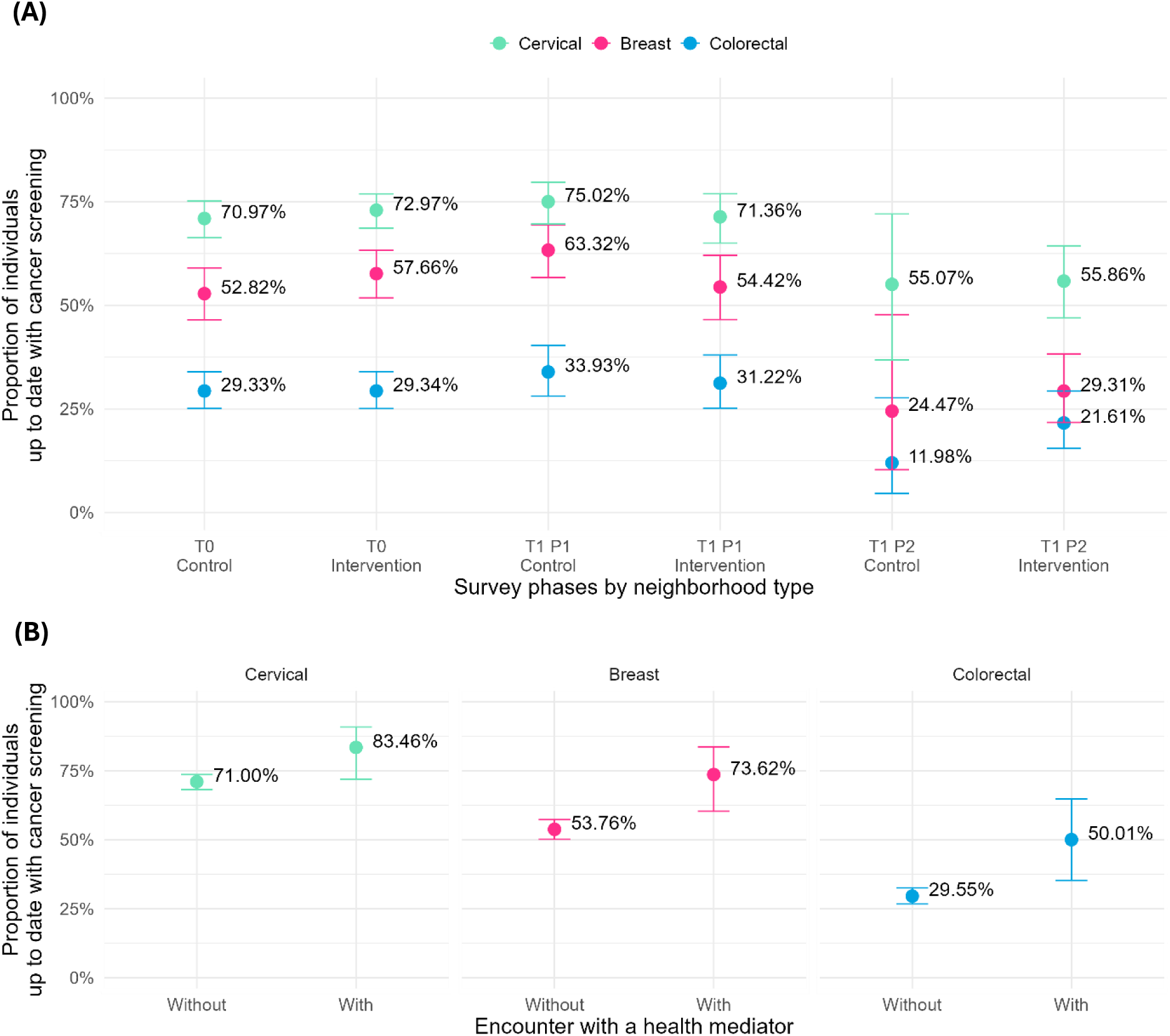
Effect of health mediation on cancer screening **(A)** at the population level and, **(B)** at the individual level.

**Figure 3.**
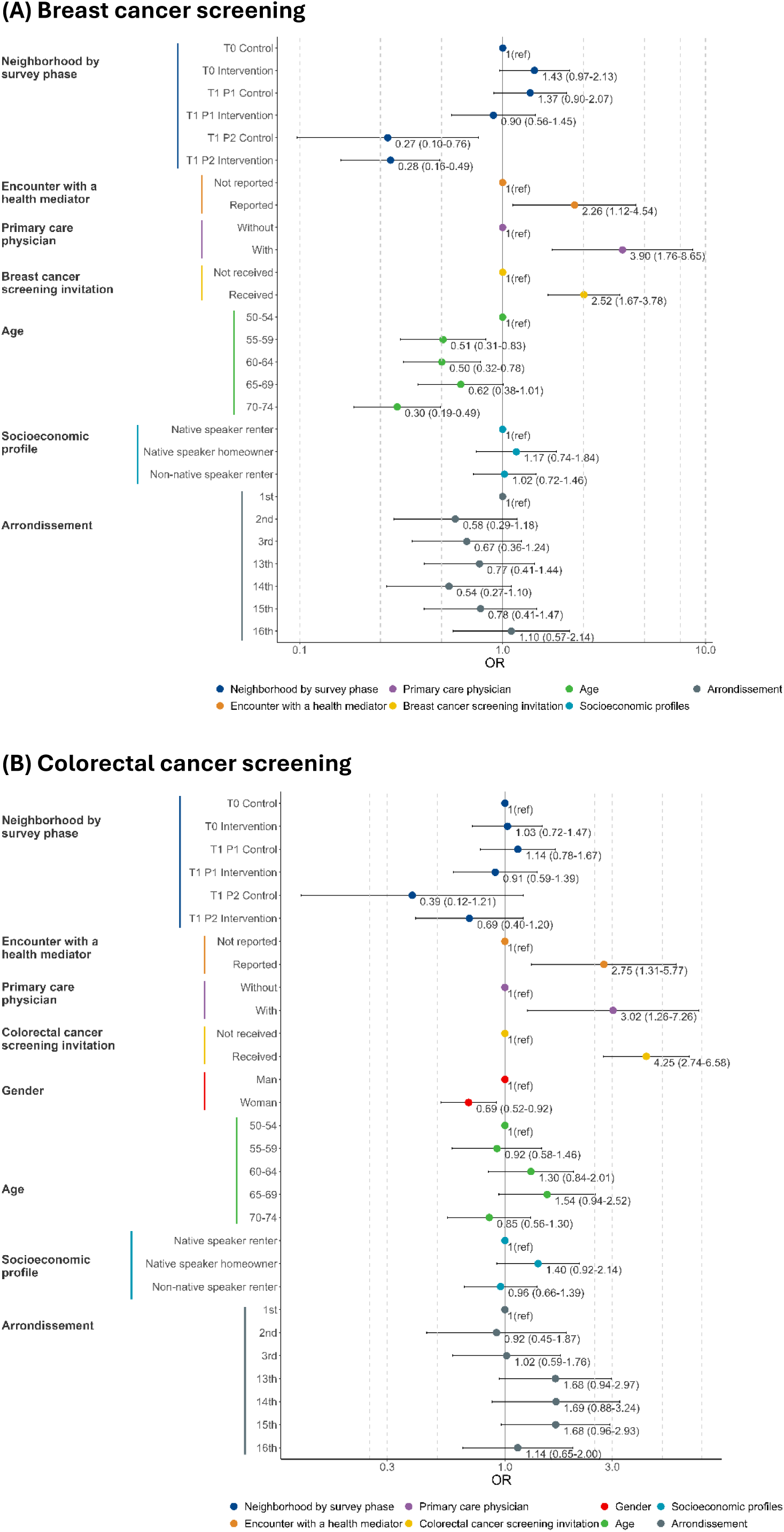
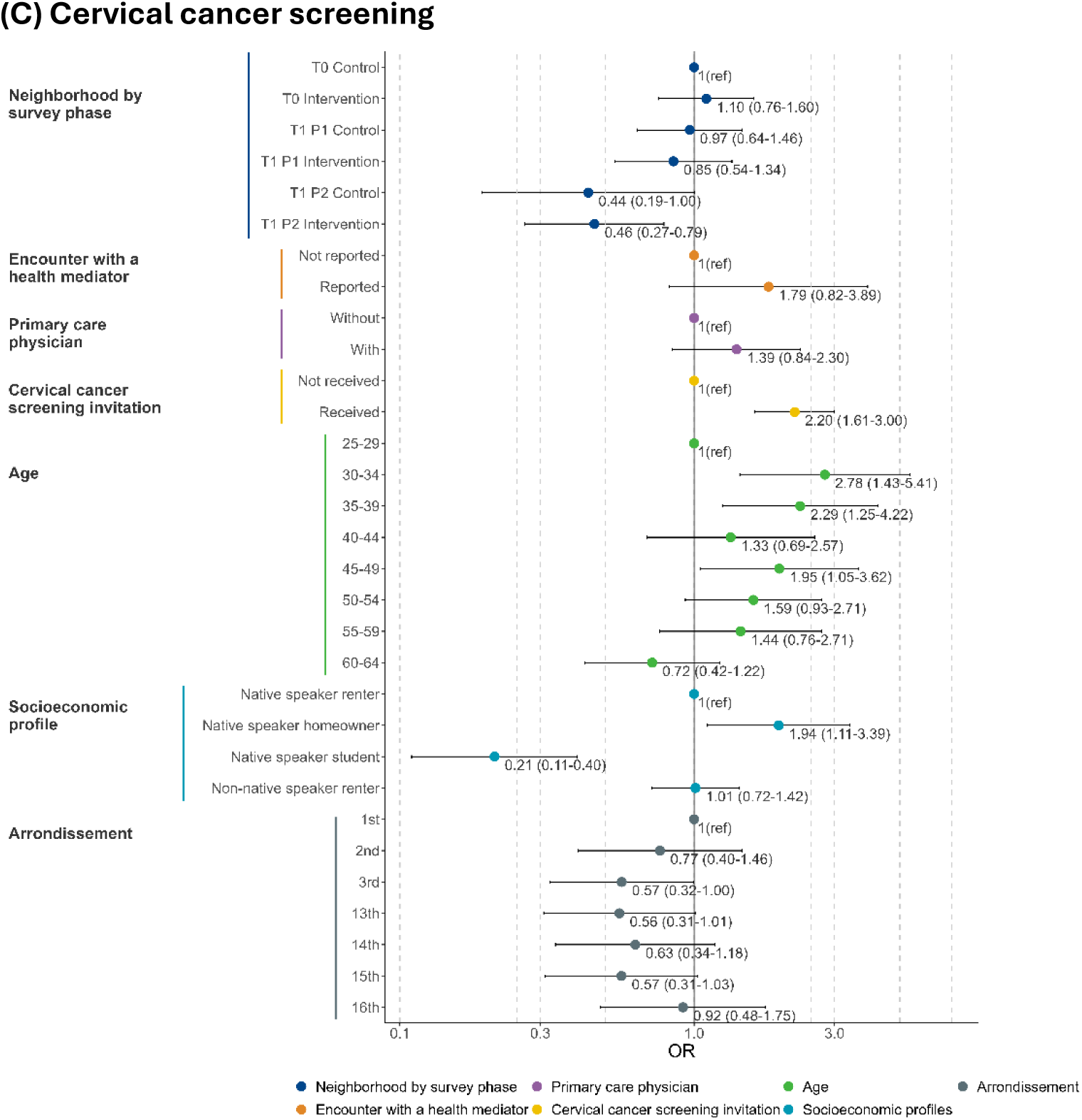
Factors associated with up-to-date cancer screening in multivariate analysis. **(A)** Breast cancer screening for women aged 50–74 years (n=1252). **(B)** Colorectal cancer screening for men and women aged 50–74 years (n=1925). **(C)** Cervical cancer screening for women aged 25–64 years (n=1666).

### Individual-level effect of health mediation on cancer screening

#### Cancer screening uptake

When exposure was defined at the individual level in the weighted multivariate analysis, participants reporting an encounter with a health mediator had significantly higher breast (WP=73.62%, n=42) and colorectal (WP=50.01%, n=43) cancer screening uptakes compared with those who had not (breast: WP=53.76%, n=679, OR=2.26 [1.12-4.54]; colorectal: WP=29.55%, n=626, OR=2.75 [1.31-5.77]) (**Figure 2B**, **Figure 3A-B, Sup. tables 7-8 p.28-29**). However, although an upward trend was observed, cervical cancer screening uptake was not significantly higher among participants who reported an encounter with a health mediator (encounter: WP=83.46%, n=72; no encounter: WP=71%, n=1319; OR=1.79 [0.82-3.89]) (**Figure 2B**, **Figure 3C, Sup. tables 9 p.30).**

Reported access to a primary care physician was also significantly associated with increased uptake in breast (with physician: WP=55.96%, n=721; without: WP=29.75%, n=17; OR=3.90 [1.76-8.65]) and colorectal cancer (with physician: WP=31.35%, n=667; without: WP=10.34%, n=10; OR=3.02 [1.26-7.26]) but not with cervical cancer screening (OR=1.39 [0.84-2.30]) (**Figure 3**, **sup. tables 7-9 p.28-30**). Only receipt of screening invitation letter significantly increased uptake for the three cancers (breast: 38.81% vs 58.83%, OR=2.52 [1.67-3.78]; colorectal: 11.92% vs 38.88%, OR=4.25 [2.74-6.58], cervical: 64.04% vs 79.05%, OR=2.20 [1.61-3.00]).

Finally, age was associated with breast and cervical cancer screening uptake. Women aged over 54 years had significantly higher uptake of breast cancer screening compared with women aged 50-54 years (OR_55-59_=0.51 [0.31-0.83], OR_60-64_=0.50 [0.32-0.78], OR_65-69_=0.62 [0.38-1.01], OR_70-74_=0.30 [0.19-0.49]) (**Figure 3A, sup. table 7 p.28**). Regarding cervical cancer screening, women aged 30–39 were more likely to report being up to date than younger women (OR_30-34_=2.78 [1.43-5.41], OR_35-39_=2.29 [1.25-4.22]) (**Figure 3C, sup. table 9 p.30**).

#### Cancer screening awareness

Participants reporting an encounter with a health mediator had significantly higher colorectal cancer screening awareness (WP=93.43%, n=86) compared with those who had not (WP=82.81%, n=1753, OR=8.07 [2.10-30.96]) (**Sup. table 10, sup. figure 16 p.31-32**). No significant effect of health mediation at individual level was observed for breast and cervical cancer screening awareness (breast: OR=3.24 [0.43-24.16], cervical: OR=2.24 [0.64-7.85]) (**sup. tables 11-12, sup. figures 17-18 p.33-36**). However, awareness of breast cancer screening was very high overall, with 96.63% of women reporting that they were aware of the national program. Overall, awareness reached 83.35% for colorectal cancer screening and 86.72% for cervical cancer screening. Non-NS renters reported lower awareness for cancer screening compared with NS renters for the three cancers (breast: OR=0.39 [0.16-0.96]; colorectal: OR=0.34 [0.21-0.54]; cervical: OR=0.26 [0.16-0.44]) (**Sup. tables 10-12, sup figures 16-18 p.31-36**).

## Discussion

This repeated cross-sectional study showed that the “*13 en Santé”* health mediation program successfully targeted individuals facing cumulative socioeconomic barriers to accessing health care (i.e., non–French native speakers, those with the lowest levels of education, and those with the highest rates of unemployment or homemaking)^23^. These individuals reported the lowest access to a primary care physician and the lowest receipt rates of cancer screening invitation letters. The reduced receipt rate may reflect an actual lower likelihood of receiving the letter — for example, due to unstable housing — or difficulties in understanding the content of the invitation. Additionally, this study shows that health mediation may significantly increase individual uptake of breast and colorectal cancer screening, with a 20-percentage-point gain. As demonstrated by the multivariate analysis, this effect may suggest a complementary role of health mediation alongside access to a primary care physician and receipt of the invitation letter.

The 20-percentage-point increase in cancer screening uptake associated with health mediation is consistent with previous studies, although slightly higher than reported median effects. A 2023 systematic review of CHW interventions effectiveness reported a median increase of 11.5 percentage points for breast cancer screening (interquartile range: 5.5–23.5) and a slightly lower median increase of 10.5 percentage points for colorectal cancer screening (interquartile range: 4.5–17.5)^13^. In contrast, despite an upward trend, no significant effect was observed in our study, although this review also reported a median increase of 12.8 percentage points (interquartile range: 6.4–21.0) for cervical cancer screening. A 2017 meta-analysis similarly demonstrated beneficial effects of community-based interventions across breast, colorectal, and cervical cancer screenings (OR=1.90 [1.60–2.26])^24^. Reported encounter with a health mediator was associated with receipt of screening invitation only for cervical cancer, which may suggest less precise targeting toward women already engaged with preventive care. Consistently, women aged 30–39 — who are more likely to have regular gynaecological follow-up related to childbearing — appeared more often up to date than other eligible women, indicating that targeting younger and older age groups may enhance intervention effectiveness.

By combing multicomponent interventions, the “13 en Santé” health mediation program may promote cancer screening through multiple mechanisms of action and by mobilizing diverse levers. First, providing tailored educational sessions or information — using motivational interviewing techniques — may enhance intrinsic motivation by addressing three fundamental needs (competence, autonomy, and relatedness) and can be further reinforced through navigation support when needed^24,25^. Second, navigation support helps address contextual barriers — whether perceived or real, social or related to accessing the healthcare system — that may hinder individuals from translating intention into action^25–27^. As a matter of fact, health mediation appeared associated with increased awareness only for colorectal cancer screening, whereas breast cancer awareness was already very high in our sample. This suggests differential mechanisms of action: for colorectal cancer, both raising awareness through tailored education and providing navigation support to overcome barriers appear necessary, while for breast cancer, the intervention may act primarily by helping individuals resolve practical barriers rather than by increasing awareness.

No effect of health mediation or primary care physician access was observed on cervical cancer screening uptake. The absence of an effect of primary care physicians on cervical cancer screening in French urban settings is consistent with litterature^28^. This finding highlights the central role of specialists — gynaecologists and midwives — in this screening pathway and underscores that its uptake driving mechanisms differ from those for breast and colorectal cancers. Further research is needed to identify the specific mechanisms that should be integrated into health mediation strategies to improve their effectiveness in promoting cervical cancer screening.

This study has a major strength in enabling assessment of the intervention through exposure defined at both the individual and neighbourhood levels, allowing simultaneous evaluation of individual- and population-level effects.

It also has several limitations. First, although the intervention was concentrated within the designated intervention neighbourhoods, it may not have been sufficient to prevent dilution effects across time and space. In the northern intervention neighbourhoods — where activities were primarily conducted through workshops and focus groups — health mediators may have interacted with individuals residing in adjacent areas, including control or non-included neighbourhoods, thereby diluting the measurable effect of the intervention. Second, although 4,523 individuals were interviewed, it represented only about 6% of the total population of the included neighbourhoods. Third, the T1 follow-up survey was conducted in two phases for operational reasons (i.e., completion of missing questionnaires in Phase 2 due to time constraints during Phase 1), which may have influenced observed screening uptakes. Although survey phase was accounted for in the statistical models, this two-phase data collection may have reduced statistical power to detect population-level effects. Combined with the potential dilution effects described above and the limited population coverage, this may have constrained the study’s ability to identify an effect at the population level. Further studies using more comprehensive medico-administrative data from the French National Health Insurance are underway to confirm these findings.

In addition, questionnaire administration relied on door-to-door visits, which aligned with the main outreach strategy in the city centre but not in the northern neighbourhoods. This discrepancy may partly explain the lower reported encounter rates in Northern areas, as interviewed individuals may have been less representative of the populations reached by the intervention, thereby potentially limiting the observed effectiveness of the program.

Other limitations are inherent to surveys based on self-reported outcomes. First, recall bias may have led to underestimation of health mediator encounters, limiting the assessment of intervention effects. Second, social desirability bias may have resulted in overestimation of screening uptake rates. Finally, a limitation of this study is inherent to observational quasi-experimental design, which does not allow for establishing a causal relationship between exposure to the intervention and increasing screenings uptake.

## Conclusion

This study provides the first evidence that a multicomponent health mediation intervention may effectively promote breast and colorectal cancer screening among underserved and under-screened populations living in socioeconomically disadvantaged urban neighbourhoods in France. These findings position health mediation as a promising strategy to reduce persistent inequalities in cancer prevention. Further research using comprehensive medico-administrative data and strengthened implementation frameworks is needed to refine intervention mechanisms, enhance scalability, and ultimately maximize its impact on screening uptake and population health.

## Abbreviations

ARS Paca: Agence Régionale de Santé Provence–Alpes–Côte d’Azur
CHW: community health worker
HPV: Human Papilloma Virus
NS: native speaker
OR: odds ratio
SpF: Santé publique France
WP: weighted percentage

## Acknowledgments

All the co-authors would like to express their deep gratitude to the health mediators of the SEPT and CORHESAN projects for their dedication, commitment, and involvement in this research—without whom this project would not have been possible—as well as for their daily work, all carried out with the aim of improving health for everyone. The co-authors also wish to thank all the members—regular or invited—of the Evaluation Technical Committee for their valuable guidance, expertise, and sustained commitment, which greatly contributed to the success of this project, with special thanks to Valérie Henry.

## Contributors

SR, FF, LR, PC, PM and JG designed the study, with contributions with DK, CG, SN, ADK and YA. SR, LR, FF, PC, PM, ADK and YA designed the methodological approach, study procedures, and tools. GB, ADK, YA, FF, LR, EL and SR supervised the operational project and collected data. EL, FF, DK and SR verified the data. EL, SR and JG designed the statistical analysis. EL and SR conducted the analysis. All authors interpreted the findings. EL created the figures and wrote the first draft of the manuscript. All authors reviewed and revised the manuscript and approved this version of the Article for publication. All authors had access to the data and accept responsibility to submit for publication.

## Data sharing

Data will be made available upon reasonable request by contacting the corresponding author.

