## Supplemental file for "Effectiveness of health mediation to promote organized cancer screening among underserved and under-screened populations in Marseille, France: findings from a repeated cross-sectional survey"

[1], Hôpital Européen, Marseille, France

[2], Prospective et Coopération, Marseille, France

[3], Association Santé Environnement pour Tous, Marseille France

[4], Agence Régionale de Santé Provence-Alpes Côte d'Azur, Marseille, France

[5], Aix Marseille Univ, IRD, INSERM, UMR1252, SESSTIM, APHM, ISSPAM, BioSTIC, Marseille, France

[6], Santé publique France Provence-Alpes-Côte d'Azur et Corse, Marseille, France

[7], SESSTIM, Aix Marseille Université, IRD, INSERM, ISSPAM, Marseille, France

### METHODS

#### APPENDIX 1. STUDY DESIGN AND SETTING

**Supplementary method 1. Health mediation teams' description**

Because of territorial specificities, CORHESAN mainly conducted door-to-door outreach visits, whereas SEPT primarily implemented workshops within partner organizations in the northern arrondissements, where door-to-door activities were unsafe, and a strong partner network was already established[1]. Both mediation teams were composed of health mediators, nurses and coordinators. Health mediators were recruited based on inter-personal skills, from different cultural origins and professional backgrounds. Every team member was trained to provide culturally adapted education or information, to conduct outreach actions with different modalities, and to help beneficiaries to navigate through healthcare systems. Even if the program was mainly focused on health, a social navigation could be carried to overcome social barriers to health.

**Supplementary method 2. Intervention and control neighborhoods selection**

Intervention neighborhoods were selected by the two operators based on their field experience and local knowledge, using criteria such as neighborhood safety and practical accessibility (e.g., areas not restricted by territorial tensions) and the strength of the local community network, to ensure feasible and rapid implementation of health mediation activities. Thirteen neighborhoods were selected: four in the northern area (one in each of the 13$^{th}$, 14$^{th}$, 15$^{th}$ and 16$^{th}$ arrondissements), representing 10,983 inhabitants, and nine in the city center (three adjacent neighborhoods within each of the 1$^{st}$, 2$^{nd}$ and 3$^{rd}$ arrondissements) covering a target population of 14,098 inhabitants.

Control neighborhoods were selected based on five criteria: non-contiguity with any intervention neighborhood to avoid action dilution; proximity to the arrondissement of the corresponding intervention neighborhood; socioeconomic similarity; and operational accessibility for the teams. Neighborhoods' socioeconomic similarity was assessed using a principal component analysis-based distance computed from census indicators, including population density, age structure, socioeconomic categories, proportions of foreign-born or immigrant residents, education level, unemployment rate, median income, proportion of manual workers, the 2015 EDI deprivation score, and housing overcrowding[2].

[1] Magnani C, de Mongolfier S, Legendre E, Rebaudet S, Mininel F. Health mediation in Marseille to promote cancer screening: settings, experiences and autonomy. *Submitted* 2025.

[2] Institut national de la statistique et des études économiques. Population en 2021. 2024. https://www.insee.fr/fr/statistiques/8268806 (accessed Dec 4, 2025).

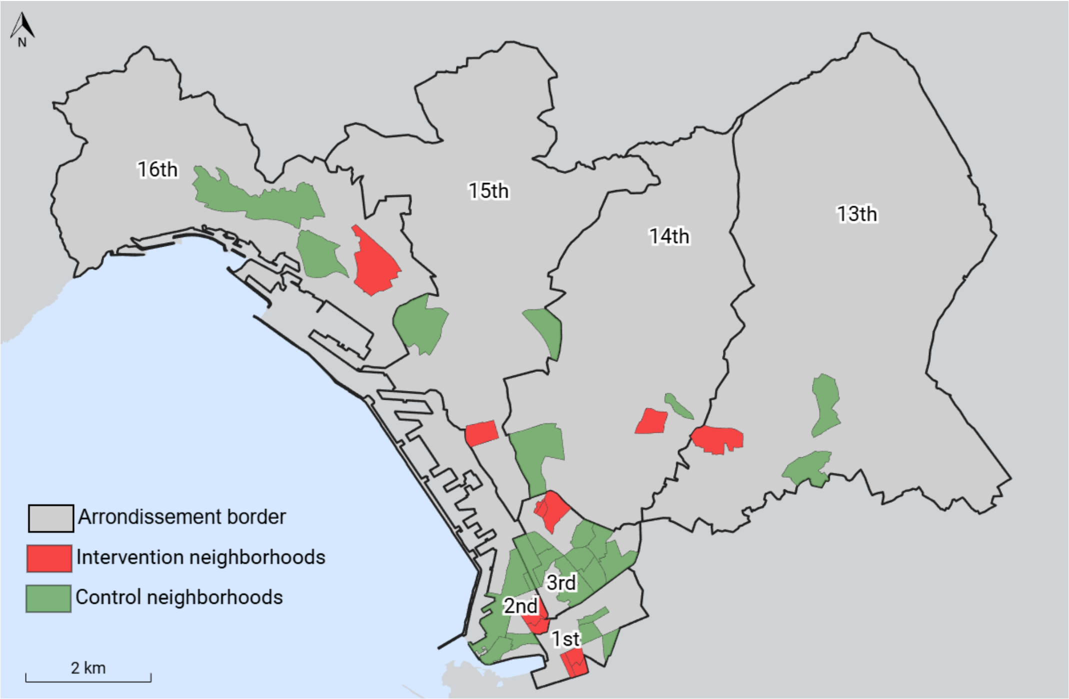

Supplementary figure 1. Intervention and control neighborhoods included in the study, located in the city-center (1st to 3rd arrondissement) and the Northern arrondissements of Marseille (13th to 16th arrondissements), France.

### APPENDIX 2. PARTICIPANTS

**Supplementary method 3. Stratification sampling method and participants' recruitment**

Participants were recruited following a predefined walking rules common to each neighborhood. The geographical starting point was randomly selected for each neighborhood. The research assistant interviewed one eligible individual per household who met the age and sex criteria, continuing until the target number of participants for each stratum (age, sex, and neighborhood) was reached.

#### Appendix 3. Procedures

Supplementary table 1. Variables' description and construction

| Variable | Modalities |
|---|---|
| **Primary outcomes: cancer screening uptake according to national guidelines** | |
| Breast cancer | ▪ Up to date: screening performed within the last two years.<br>▪ Not up to date: never screened or last screening more than two years ago. |
| Colorectal cancer | ▪ Up to date: screening performed within the last two years.<br>▪ Not up to date: never screened or last screening more than two years ago. |
| Cervical cancer | ▪ Up to date: screening performed within the past 1 year for women aged 25–26, within the past 3 years for women aged 27–30, and within the past 5 years for women aged 30–64.<br>▪ Not up to date: never screened or last screening performed outside the recommended interval for the corresponding age group. |
| **Secondary outcomes: cancer screening awareness** | |
| Breast cancer | ▪ Aware: has ever heard of breast cancer screening.<br>▪ Not aware: never heard of breast cancer screening. |
| Colorectal cancer | ▪ Aware: has ever heard of colorectal cancer screening.<br>▪ Not aware: never heard of colorectal cancer screening. |
| Cervical cancer | ▪ Aware: has ever heard of breast cancer screening.<br>▪ Not aware: never heard of cervical cancer screening. |
| **Health mediation** | |
| Encounter with a health mediator | ▪ Reported: has met health mediators from the SEPT or CORHESAN since September 2022 to discuss cancer screening and vaccination.<br>▪ Not reported: has not met health mediators from the SEPT or CORHESAN since September 2022 to discuss cancer screening and vaccination. |
| **Access to healthcare system** | |
| Primary care physician | ▪ With: has a regular primary care physician<br>▪ Without: does not have a regular primary care physician |
| Breast cancer screening invitation | ▪ Received: has been invited by her physician or by a letter from the National Health Insurance<br>▪ Not received: has not been invited by her physician or by a letter from the National Health Insurance |
| Colorectal cancer screening invitation | ▪ Received: has been invited by her physician or by a letter from the National Health Insurance<br>▪ Not received: has not been invited by her physician or by a letter from the National Health Insurance |
| Cervical cancer screening invitation | ▪ Received: has been invited by her physician or by a letter from the National Health Insurance<br>▪ Not received: has not been invited by her physician or by a letter from the National Health Insurance |
| **Socioeconomic characteristics** | |
| Birthplace | ▪ France: born in France ("Overseas Departments and Territories" and "Metropolitan France" combined due to low number of observations).<br>▪ Abroad: born abroad. |
| Mother tongue | ▪ French: French mother tongue<br>▪ Other: non-French mother tongue |
| Education level | ▪ Primary school: first stage of compulsory education, including preschool and elementary school for children aged 3 to 11 or equivalent |

| | |
|---|---|
| | ▪ <u>Secondary school</u>: second stage of compulsory education for children aged 12 to 18, including high school or equivalent.<br>▪ <u>Undergraduate</u>: first level of higher education, pursuing a bachelor's degree or equivalent.<br>▪ <u>Postgraduate</u>: advanced studies, such as a master's, doctoral (PhD), or professional postgraduate qualification.<br>▪ <u>Other</u>: all other form of education. |
| Occupational status | ▪ <u>Employed</u>: currently working for pay or profit, either full-time, part-time, or self-employed.<br>▪ <u>Student</u>: enrolled in an educational program, combined with paid apprentice/intern due to the low number of observations (similarity validated by multiple correspondence analysis).<br>▪ <u>Homemaker</u>: manages and cares for their household full-time, without formal employment.<br>▪ <u>Retired</u><br>▪ <u>Unemployed</u>: not currently working for pay or profit.<br>▪ <u>Other</u>: all other form of occupational status. |
| Residential status | ▪ <u>Homeowner</u>: individual who owns the property they live in.<br>▪ <u>Renter</u>: individual who rent the property they live in.<br>▪ <u>Family</u>: individual living in the family household.<br>▪ <u>Other</u>: combined 'Friend' and 'squat/shelter' with all other form of residential status due to low number of observations |
| Socioeconomic profile | Profiles constructed using a multiple correspondence analysis based on socioeconomic characteristics, followed by hierarchical agglomerative clustering. Four profiles were identified and labeled based on their predominant characteristics:<br>▪ Native speaker homeowner (NS homeowner)<br>▪ Native speaker renter (NS renter)<br>▪ Native speaker student (NS student)<br>▪ Non-native speaker renter (Non-NS renter)<br>See pages 9 to 14 for more details. |
| **Demographic characteristics** | |
| Gender | Self-reported gender: <u>female</u> or <u>male</u>. Missing data corresponded to a participant declaring undifferentiated gender, but due to low total headcount and statistical limit, we couldn't consider this category in our analysis |
| Age | Self-reported age was collected as a quantitative variable and then discretized into 5-year intervals. |
| Arrondissement | District of the residential neighborhood. |

### RESULTS

#### APPENDIX 4. SOCIOECONOMIC PROFILES CONSTRUCTION AND DESCRIPTION

##### 4.1. Socioeconomic profiles construction

The distribution of individuals on the first two axes of the multiple correspondence analysis, which together accounted for 25·97% of the total variance (15·36% for the first axis and 10·61% for the second), showed clear discrimination according to mother tongue (French vs other), place of birth (France vs abroad) — two highly correlated variables — as well as residential and occupational status. Living in the family home was strongly associated with being a student.

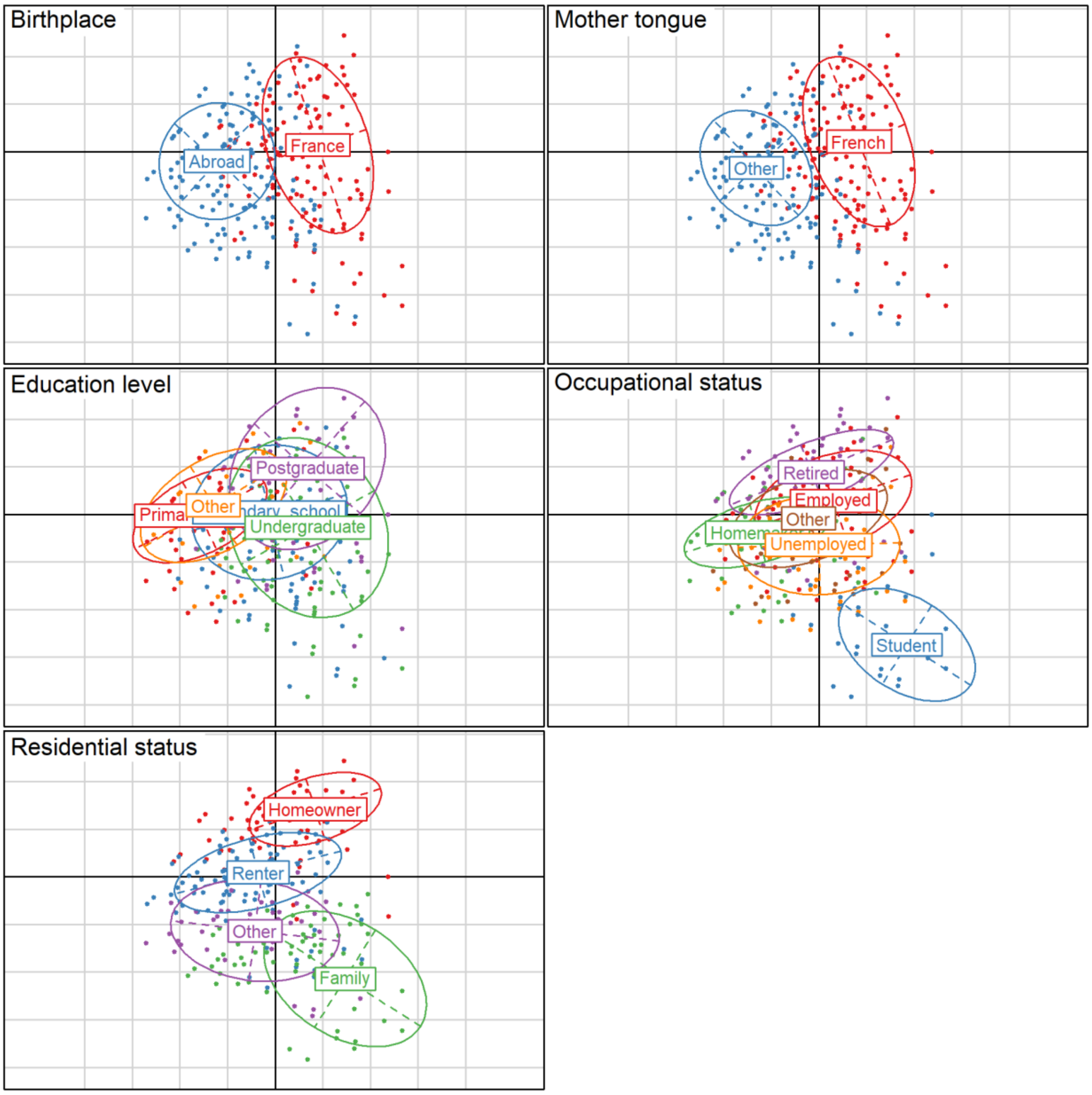

Supplementary figure 2. Distribution of individuals on the first two axes of the multiple correspondence analysis by variable categories.

A multiple correspondence analysis was performed on socioeconomic characteristics, followed by hierarchical agglomerative clustering to identify major socioeconomic profiles. Each cluster represents a group of individuals who are as similar as possible in terms of their socioeconomic characteristics while being distinct from other groups. These clusters provide a statistical approximation of social patterns within the population and are useful for identifying groups accumulating multiple social disadvantages, although some heterogeneity within clusters inevitably remains.

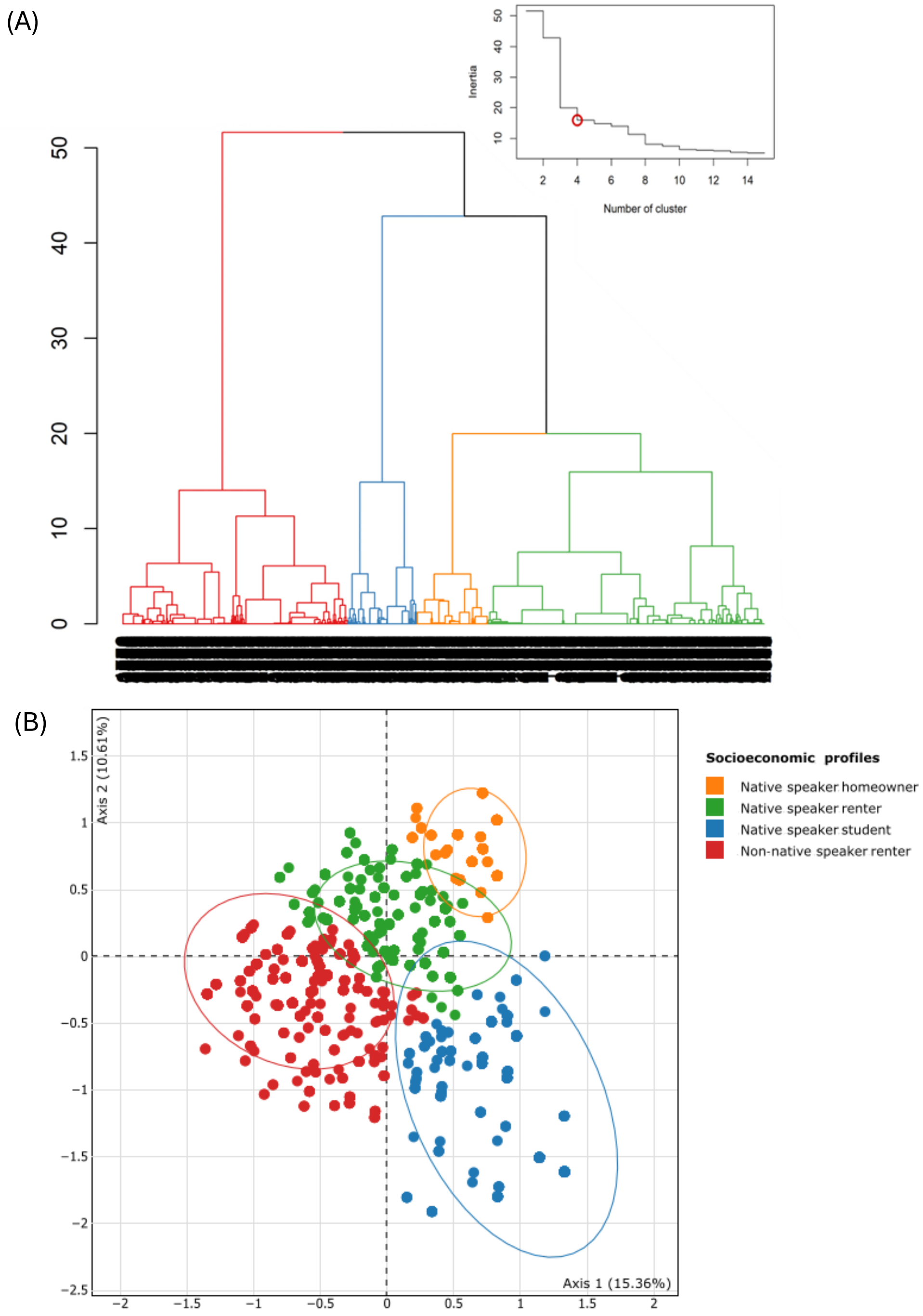

Supplementary figure 3. Hierarchical agglomerative clustering after multiple correspondence analysis. (A) Dendrogram and inertia. (B) Distribution of individuals on the first two axes of the multiple correspondence analysis by the four socioeconomic profiles identified by the clustering.

#### 4.2. Socioeconomic profiles description before imputation

Four socioeconomic profiles were identified and named according to the most discriminating characteristics within each group: native speaker homeowner (n=483; 11·14%), native speaker renter (n=1862; 42·96%), native speaker student (n=467; 10·78%), and non-native speaker renter (n=1522; 35·12%). A total of 189 individuals out of 4523 participants (4·18%) had missing socioeconomic profiles.

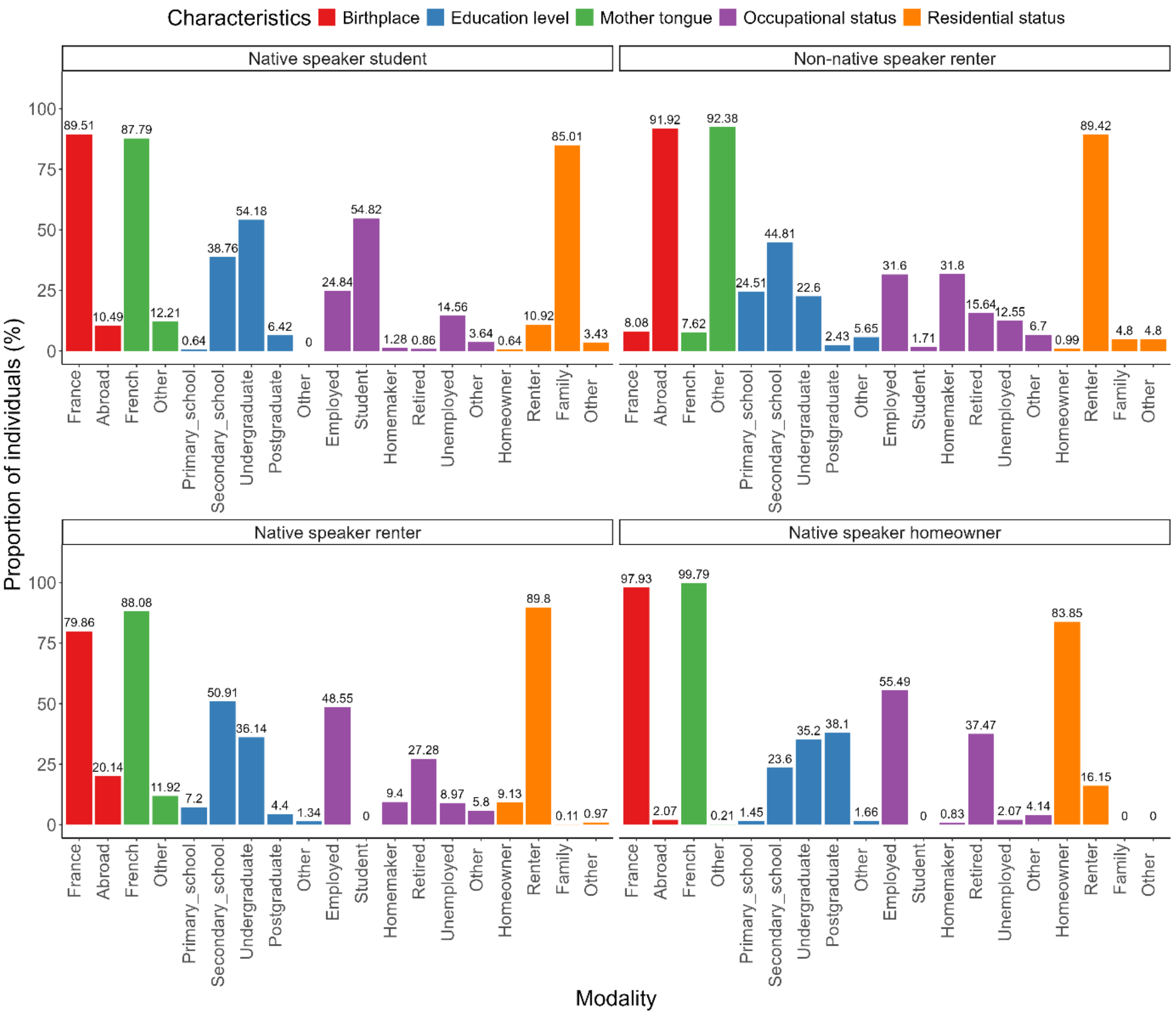

Supplementary figure 4. Socioeconomic characteristics of participants in the four profiles identified by clustering before missing data imputation.

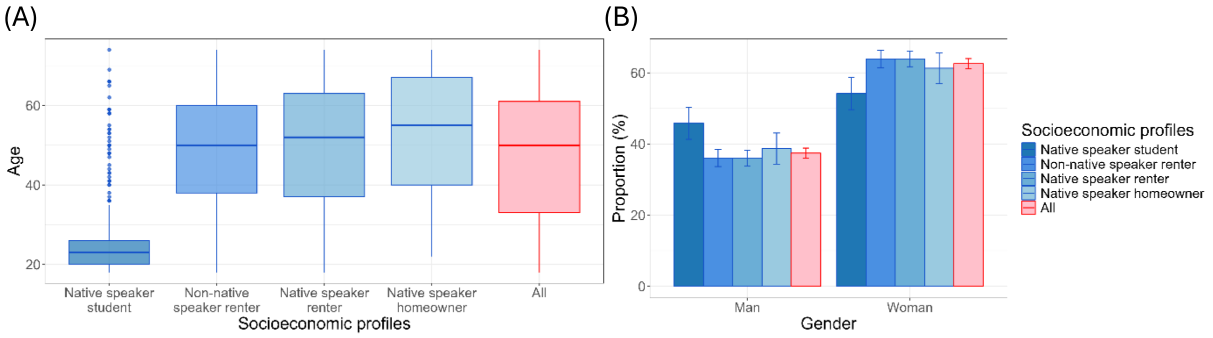

Supplementary figure 5. (A) Age and (B) gender distribution across the four socioeconomic profiles.

Among the four profiles, the "non-native speaker renter" profile was distinct from the others, as it included 92% of participants born abroad and without French as their mother tongue. The other three groups mostly comprised French native speakers born in France. This profile was also characterized by a high proportion of renters (90%) and the highest proportion of individuals with only primary education (25% vs <8% in the other groups). It also included the highest proportion of homemakers, comprising 32% of individuals.

The "native speaker student" profile was characterized by 55% students and 85% of participants living with their family. Educational attainment ranged from secondary to undergraduate level, with the highest proportion of undergraduates. Individuals in this profile had a substantially lower median age compared with the other groups (23 years vs 50–55 years), suggesting that many were likely still in education, which may explain the lower proportion of postgraduates. The proportion of men in this group was also higher than in the overall population (46% vs 37%).

The "native speaker homeowner" profile was characterized by the highest proportion of homeowners, comprising 84% of the group compared with <10% in the other profiles, and the highest proportion of postgraduates (38% vs <7% in the other groups). This profile also had the highest median age (55 years), with over 20% of participants aged 70–74 years (21·74% vs <11% in the other groups), resulting in the highest proportion of retirees. It also included one of the highest proportions of employees (55%).

Finally, the "native speaker renter" profile, representing 43% of the surveyed population, included French native speakers, 90% of whom were renters. This group also had one of the highest proportions of employees (49%).

#### 4.3. Socioeconomic profile imputation for missing data

Overall, at least one socioeconomic variable (birthplace, education level, occupational status, mother tongue, or residential status) was missing for 189 (4·18%) of 4523 participants. Occupational status accounted for the highest proportion of missing data (n=94; 2·08%; table 1). Socioeconomic cluster imputation was applied to these 189 participants. Imputation was performed using predicted missing clusters derived from a regression tree constructed with individuals who had complete socioeconomic information, from which their socioeconomic cluster could be assigned. Supplementary figure 6 displayed the regression tree. According to the regression tree, when no socioeconomic information was available, the cluster "native speaker renter" was imputed, as it was the most frequent in the study population (n=1862; 42·96%).

However, because this cluster represented only 43% of the population, the probability of misclassification was approximately 67%. Therefore, when no data were available, no cluster was assigned.

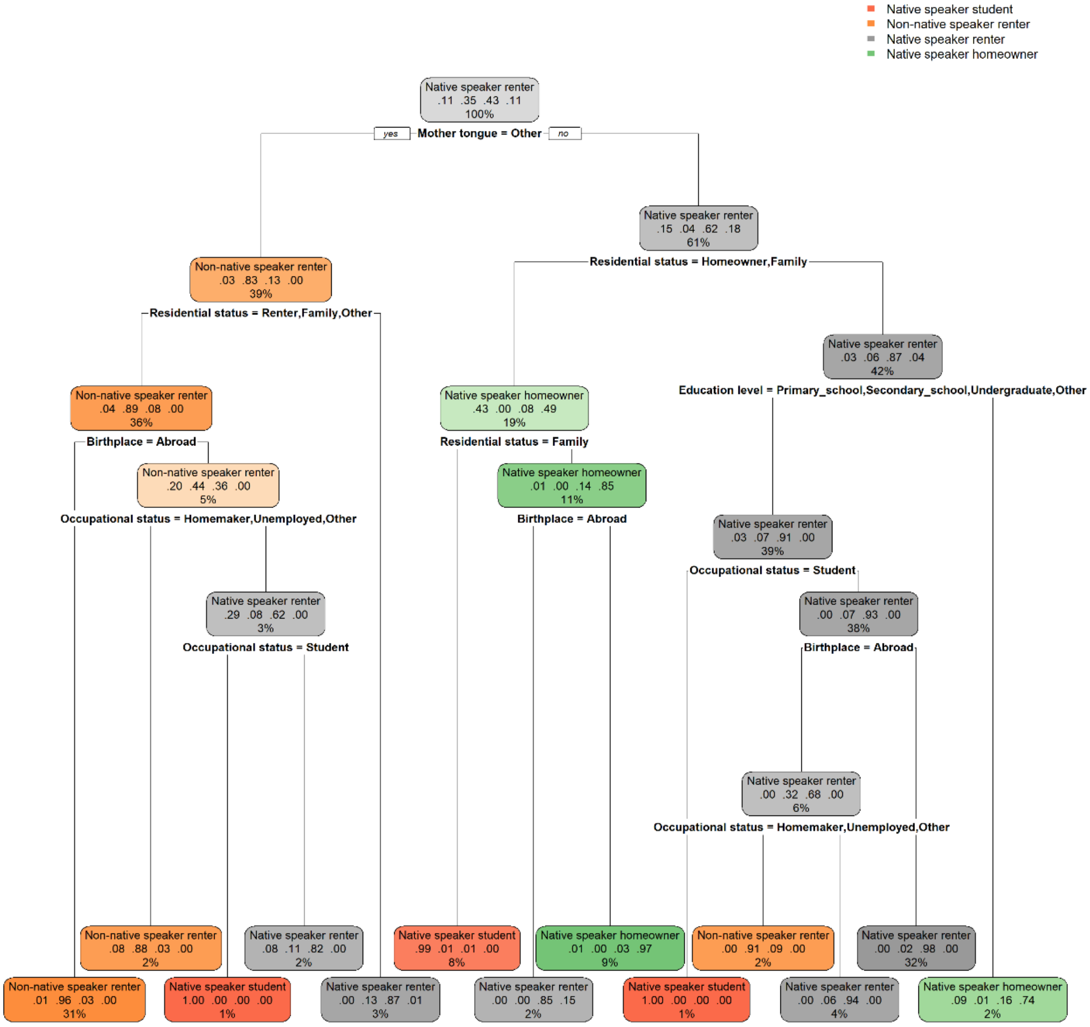

Supplementary figure 6. Regression tree of profiles based on socioeconomic variables (n=4334).
Each node of the regression tree is represented by a rectangle. Within each rectangle, the proportion of the total population in the node and its distribution across the four socioeconomic clusters (native speaker student, non-native speaker renter, native speaker renter, and native speaker homeowner, in this order) are displayed. The socioeconomic cluster labelling the node corresponds to the class assigned. For example, the first node of the tree is labelled “native speaker renter.” When no socioeconomic information was available for a participant, the class “native speaker renter” was imputed, as it was the most frequent class in the study population (n=1862, 42·96%). The variable used to partition the sample is shown below each node. For example, the first partition was based on the variable “mother tongue,” with the category “Other” appearing after the “=”. On the left side of the tree, participants with a mother tongue other than French are grouped, whereas on the right side, participants with French as their mother tongue are grouped.
In the second right node, participants with French as their mother tongue accounted for 61% (2648 of 4334 participants) of the population. In the absence of additional socioeconomic information, the “native speaker renter” cluster was assigned, as it was the most frequent class in this subgroup (1640 of 2648 participants, 61·93%). This group was then divided according to residential status. Participants who reported being home owner or living in family home were grouped in the left node and all assigned to the “native speaker student” cluster, whereas those who did not report being students were grouped in the right node.

#### 4.4. Socioeconomic profiles description after imputation

The four socioeconomic profiles remained unchanged after missing data imputation, and their distributions were largely stable: native speaker homeowner (n=493, 10·90% vs n=483, 11·14% before imputation), native speaker renter (n=1938, 42·85% vs n=1862, 42·96% before imputation), native speaker student (n=477, 10·55% vs n=467, 10·78% before imputation), and non-native speaker renter (n=1608, 35·55% vs n=1522, 35·12% before imputation). Seven individuals (0·15%) still had missing socioeconomic profiles because all socioeconomic variables were missing.

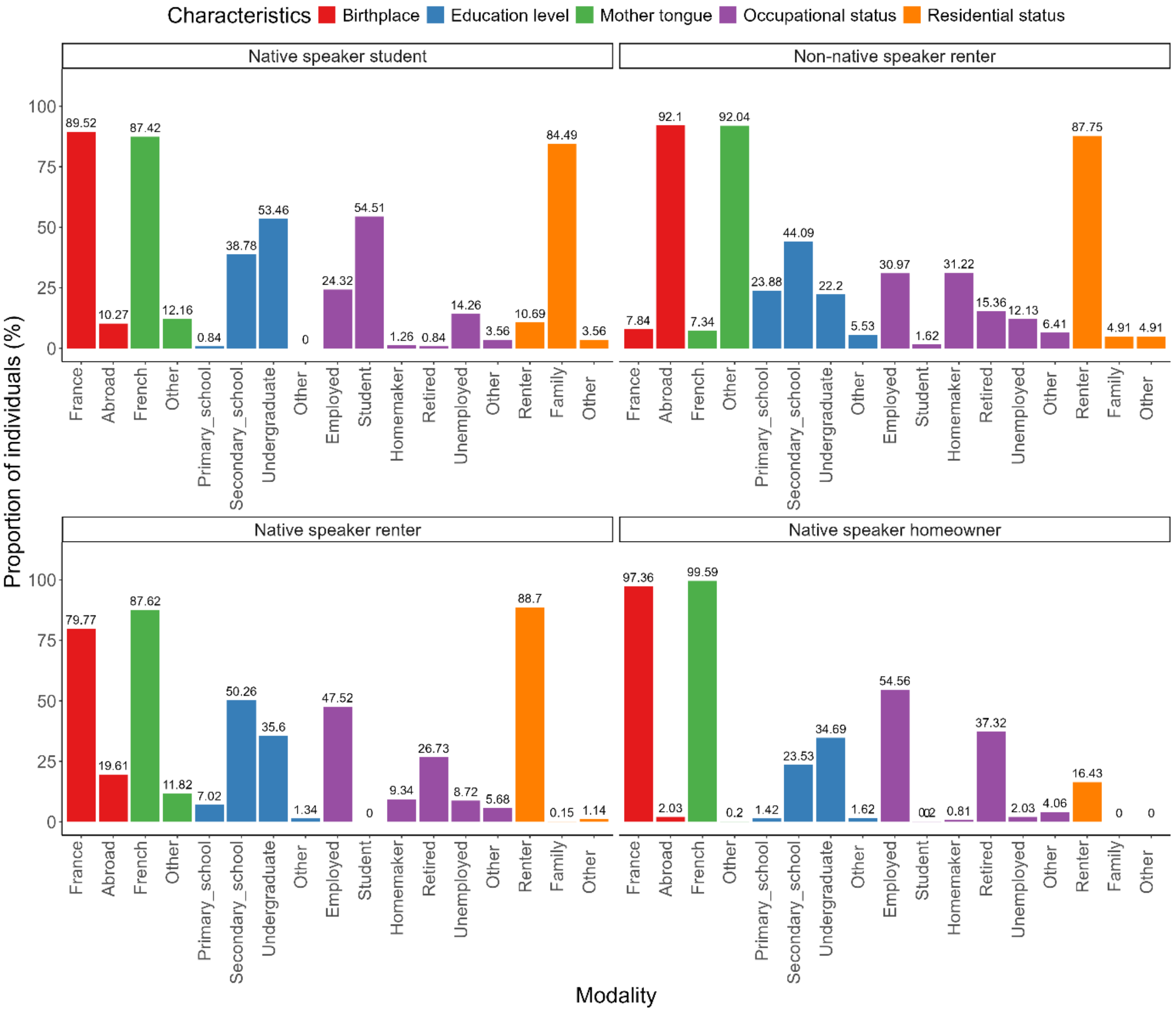

Supplementary figure 7. Socioeconomic characteristics of participants in the four profiles identified by clustering after missing data imputation.

### APPENDIX 5. IMBALANCE BETWEEN GENERAL POPULATION AND THE SAMPLE SURVEYED AND WEIGHTING

Because of the sampling design, women and individuals aged 65–74 years were overrepresented in the surveyed sample (Supplementary figure 8). To adjust the sample to the structure of the general population by age and sex, higher weights were assigned to men and to younger age groups (Supplementary figure 9). This weighting procedure reduced discrepancies with the general population and produced a sample structure representative of the general population in terms of age and sex (Supplementary figure 10).

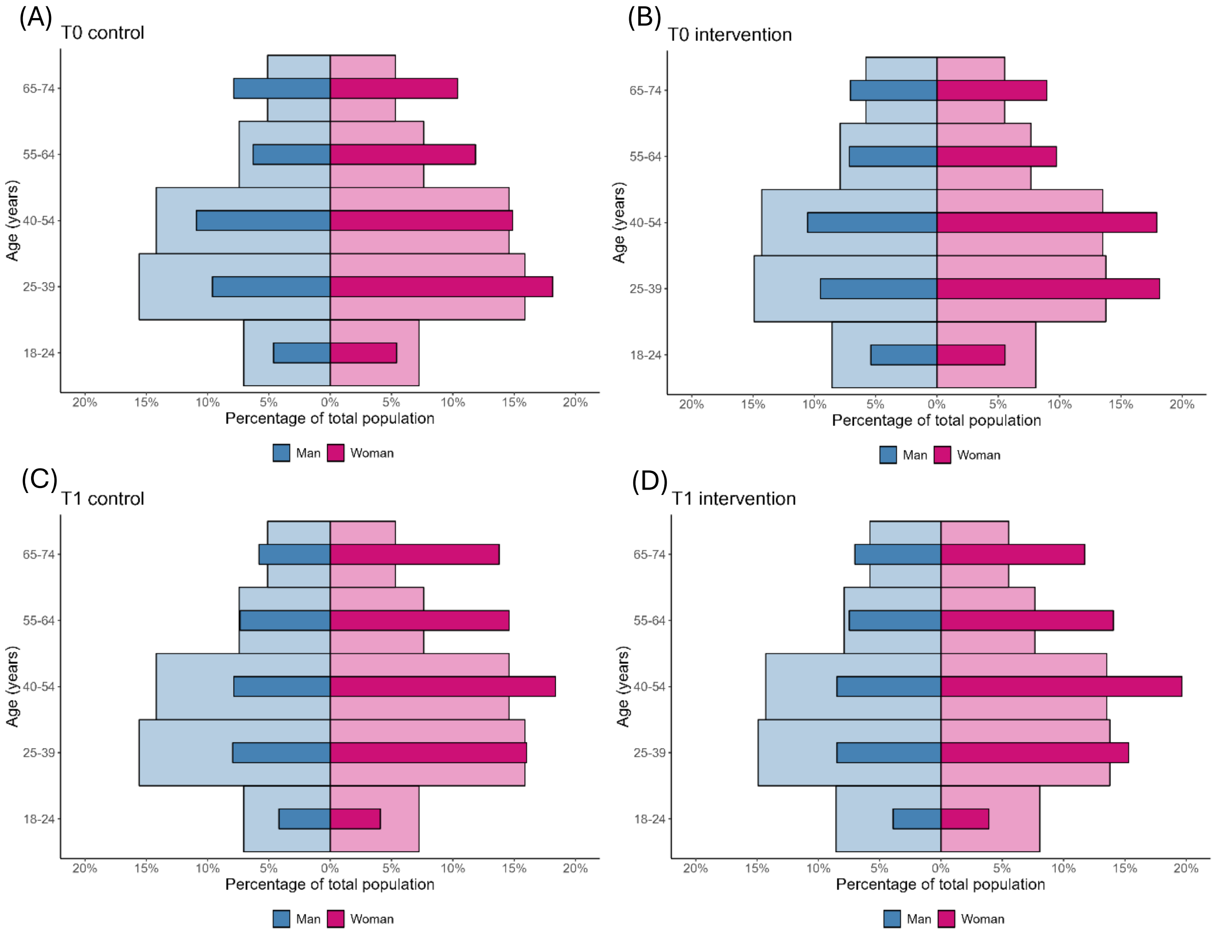

Supplementary figure 8. Comparison of age pyramids for the general population (wide light-colored bars) and the survey samples (narrow dark-colored bars) in (A) T0 control, (B) T0 intervention, (C) T1 control, and (D) T1 intervention neighborhoods.

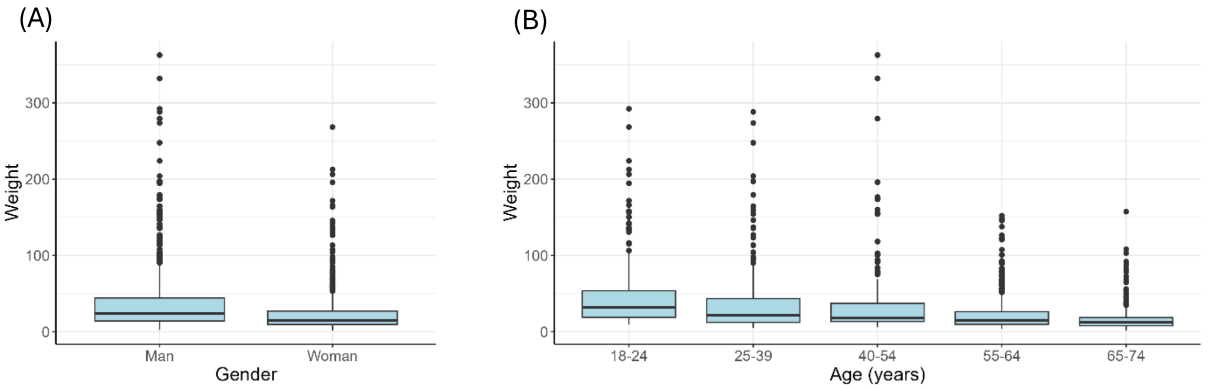

Supplementary figure 9. Weight distribution of the surveyed sample according to (A) gender and (B) age.

Although socioeconomic profiles were not directly included in the weighting process, this adjustment also affected their distribution within the sample. Specifically, it increased the relative weight of younger participants, particularly students, as observed in Supplementary figure 5 (Supplementary figure 11). A significant difference remained between the proportions of "non-native speaker renter" profiles at T0 and T1 in intervention neighborhoods (T0, 41·51% [38·31–44·78] vs T1, 31·19% [27·71–34·90]). The socioeconomic profile should therefore be considered as a potential confounding factor in the effectiveness models.

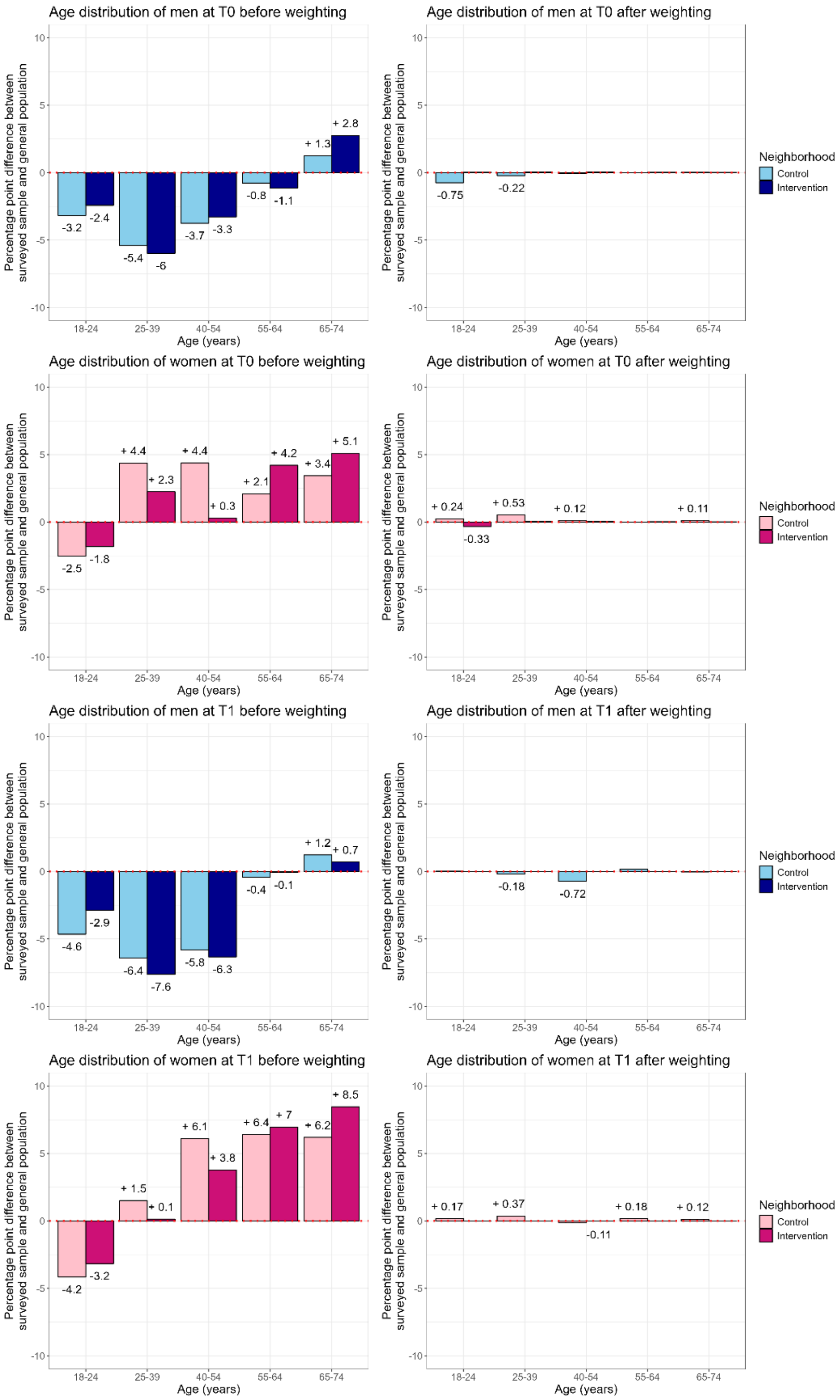

Supplementary figure 10. Percentage point difference of age and gender distributions between surveyed sample and general population, according to neighborhood type and survey time, before and after weighting.

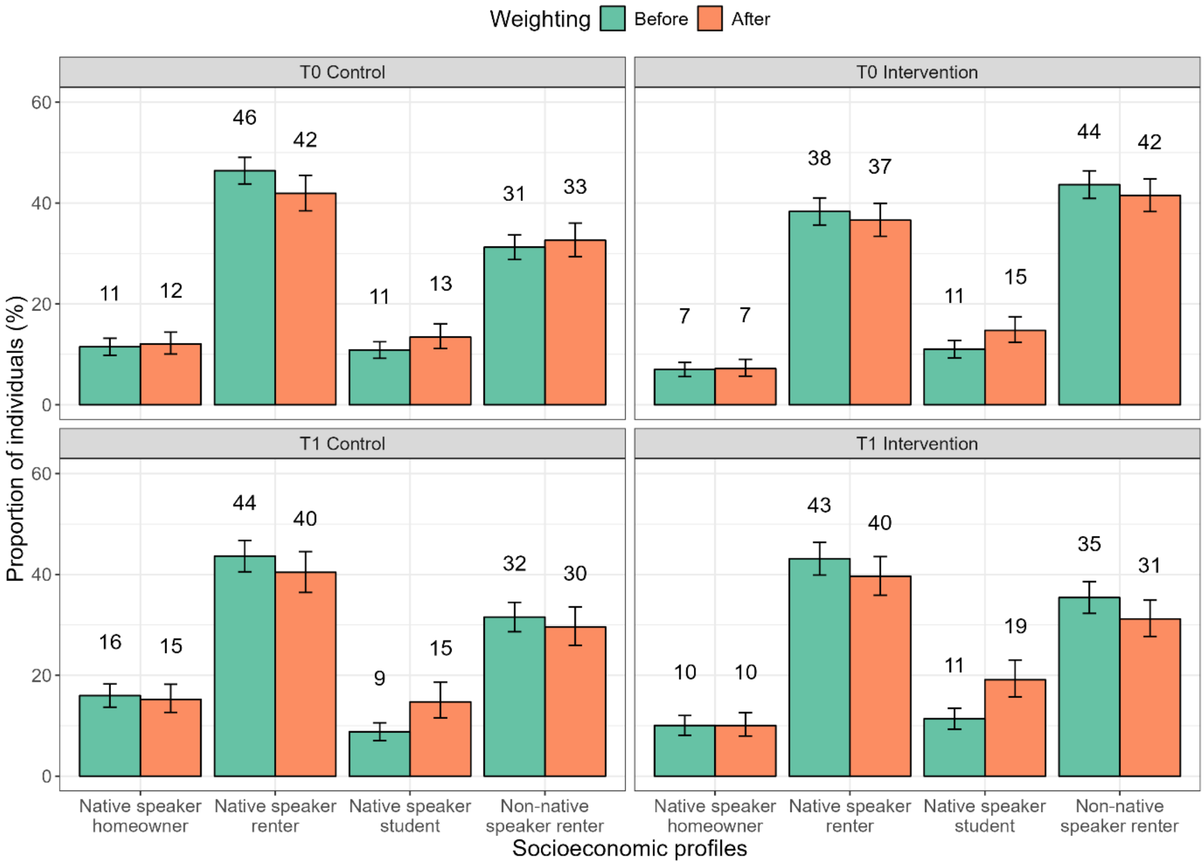

Supplementary figure 11. Proportion of socioeconomic profiles in the surveyed sample before and after weighting, according to neighborhood type and survey time.

### APPENDIX 6. ACCESS TO HEALTHCARE SYSTEM

#### 6.1. Primary care physician

Participants surveyed at T1 (T1 P1 control: OR = 2.11 [1.43-3.10], T1 P1 intervention: OR = 3.12 [1.82-5.33], T1 P2 control: OR = 3.70 (1.17-11.75), women (OR = 1.91 [1.45-2.51]) and participants aged over 40 years (from 40-44 years : OR = 3.15 [1.52-6.53] to 70-74 years : OR = 4.73 [2.00-11.18]) declared a higher access primary care to physician than participants of the T0 survey, men and those aged lower than 40 years, respectively.

Supplementary table 2. Factors associated with individuals with a primary care physician.

| | n (%) | N with physician (weighted %) | Univariate analysis Odds ratio (95% CI) | p value | Multivariate analysis Odds ratio (95% CI) | p value |
|---|---|---|---|---|---|---|
| **Primary care physician** (n [with primary care physician] = 4 000 (WP = 85.14%); n [without missing outcome] = 4 509; n [multivariable analysis] =4 401) | | | | | | |
| **Neighborhoods by survey phase** | | | | | | |
| T0 Control | 1379 (30.58) | 1183 (80.63) | 1 (ref) | .. | 1 (ref) | .. |
| T0 Intervention | 1260 (27.94) | 1085 (82.61) | 1.14 (0.88-1.48) | 0.32 | 1.12 (0.84-1.50) | 0.42 |
| T1 P1 Control | 884 (19.61) | 806 (88.98) | 1.94 (1.37-2.74) | 0.0002 | 2.11 (1.43-3.10) | 0.0002 |
| T1 P1 Intervention | 580 (12.86) | 550 (92.77) | 3.08 (1.92-4.95) | <0.0001 | 3.12 (1.82-5.33) | <0.0001 |
| T1 P2 Control | 81 (1.80) | 77 (91.11) | 2.46 (0.86-7.00) | 0.09 | 3.70 (1.17-11.75) | 0.03 |
| T1 P2 Intervention | 281 (6.23) | 260 (88.58) | 1.86 (1.07-3.23) | 0.03 | 1.73 (0.94-3.19) | 0.08 |
| Missing data | 44 (0.98) | 39 (89.15) | .. | .. | .. | .. |
| **Encounter with a health mediator** | | | | | | |
| Not reported | 4280 (94.92) | 3785 (85.25) | 1 (ref) | .. | | |
| Reported | 162 (3.59) | 155 (95.03) | 3.31 (1.28-8.57) | 0.0137 | 1.82 (0.61-5.44) | 0.28 |
| Missing data | 67 (1.49) | 60 (83.01) | .. | .. | | |
| **Gender** | | | | | | |
| Man | 1681 (37.28) | 1401 (81.79) | 1 (ref) | .. | 1 (ref) | .. |
| Woman | 2828 (62.72) | 2599 (89.11) | 1.82 (1.42-2.34) | <0.0001 | 1.91 (1.45-2.51) | <0.0001 |
| **Age** | | | | | | |
| 18-24 | 427 (9.47) | 339 (78.85) | 1 (ref) | .. | 1 (ref) | .. |
| 25-29 | 436 (9.67) | 329 (71.81) | 0.68 (0.45-1.05) | 0.08 | 0.64 (0.39-1.04) | 0.07 |
| 30-34 | 351 (7.78) | 287 (79.73) | 1.06 (0.65-1.72) | 0.83 | 1.01 (0.57-1.82) | 0.96 |
| 35-39 | 391 (8.67) | 334 (81.51) | 1.18 (0.73-1.93) | 0.50 | 1.09 (0.60-1.99) | 0.77 |
| 40-44 | 301 (6.68) | 276 (91.20) | 2.78 (1.52-5.09) | 0.0009 | 3.15 (1.52-6.53) | 0.0020 |
| 45-49 | 325 (7.21) | 295 (89.05) | 2.18 (1.24-3.84) | 0.0070 | 2.22 (1.17-4.21) | 0.01 |
| 50-54 | 593 (13.15) | 536 (89.37) | 2.26 (1.41-3.61) | 0.0007 | 1.89 (1.04-3.42) | 0.04 |
| 55-59 | 439 (9.74) | 408 (93.82) | 4.07 (2.30-7.23) | <0.0001 | 3.80 (1.94-7.44) | 0.0001 |
| 60-64 | 433 (9.60) | 412 (94.47) | 4.58 (2.03-10.35) | 0.0003 | 3.92 (1.64-9.37) | 0.0021 |
| 65-69 | 372 (8.25) | 359 (96.28) | 6.94 (3.35-14.40) | <0.0001 | 5.73 (2.57-12.79) | <0.0001 |
| 70-74 | 441 (9.78) | 425 (95.19) | 5.31 (2.49-11.34) | <0.0001 | 4.73 (2.00-11.18) | 0.0004 |
| **Socioeconomic profile** | | | | | | |
| NS renter | 1934 (42.89) | 1764 (88.81) | 1 (ref) | .. | 1 (ref) | .. |
| NS homeowner | 492 (10.91) | 452 (92.00) | 1.45 (0.93-2.27) | 0.10 | 1.42 (0.85-2.38) | 0.18 |
| NS student | 476 (10.56) | 379 (78.66) | 0.46 (0.32-0.68) | 0.0001 | 0.71 (0.44-1.16) | 0.17 |
| Non-NS renter | 1605 (35.60) | 1403 (82.07) | 0.58 (0.43-0.77) | 0.0002 | 0.58 (0.42-0.79) | 0.0007 |
| Missing data | 2 (0.04) | 2 (100.00) | .. | .. | .. | .. |
| **Arrondissement** | | | | | | |
| 1st | 531 (11.78) | 437 (80.26) | 1 (ref) | .. | 1 (ref) | .. |
| 2nd | 540 (11.98) | 474 (88.02) | 1.81 (1.17-2.80) | 0.0080 | 1.84 (1.12-3.01) | 0.02 |
| 3rd | 1192 (26.44) | 1013 (82.13) | 1.13 (0.79-1.61) | 0.50 | 1.22 (0.80-1.86) | 0.36 |
| 13th | 695 (15.41) | 658 (93.67) | 3.64 (2.20-6.02) | <0.0001 | 3.80 (2.19-6.60) | <0.0001 |
| 14th | 395 (8.76) | 353 (87.82) | 1.77 (1.12-2.81) | 0.01 | 1.83 (1.10-3.03) | 0.02 |
| 15th | 663 (14.70) | 614 (90.83) | 2.44 (1.56-3.80) | 0.0001 | 2.83 (1.72-4.66) | <0.0001 |
| 16th | 493 (10.93) | 451 (90.67) | 2.39 (1.52-3.76) | 0.0002 | 2.00 (1.19-3.34) | 0.0085 |

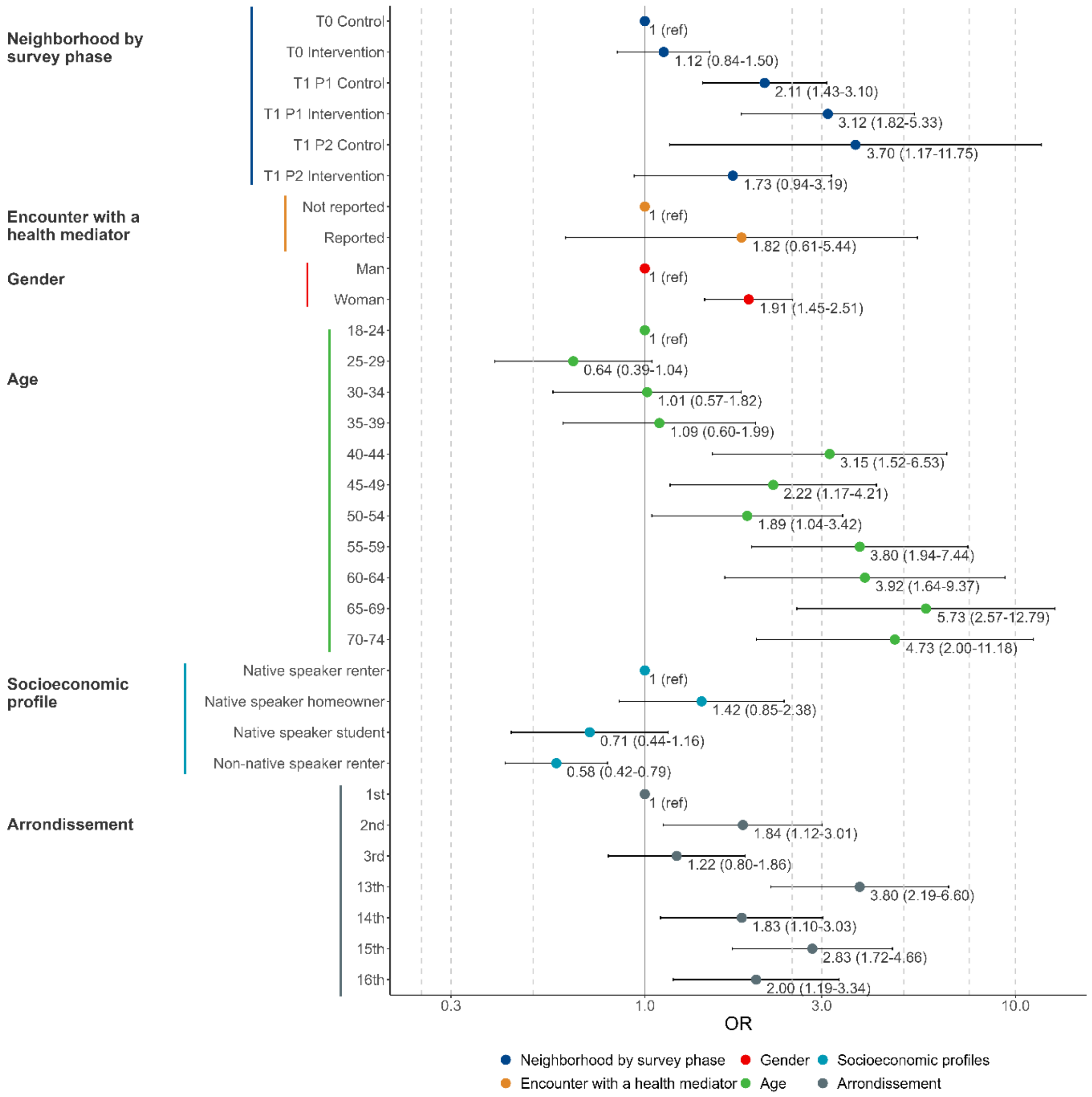

Supplementary figure 12. Factors associated with individuals with a primary care physician, in the weighted multivariate analysis (n=4 401).

#### 6.2. Breast cancer screening invitation

Supplementary table 3. Factors associated with reception of breast cancer screening invitation.

| | n (%) | n receipt invitation (weighted %) | Univariate analysis Odds ratio (95% CI) | p value | Multivariate analysis Odds ratio (95% CI) | p value |
|---|---|---|---|---|---|---|
| **Breast cancer screening invitation reception** (women aged 50–74 years; n [receipt invitation] = 1140 (WP=80.97%); n [without missing outcome] =1334; n [multivariate analysis] = 1284) | | | | | | |
| **Neighborhoods by survey phase** | | | | | | |
| T0 Control | 363 (27.21) | 318 (87.32) | 1 (ref) | .. | 1 (ref) | .. |
| T0 Intervention | 297 (22.26) | 248 (82.47) | 0.68 (0.42-1.12) | 0.13 | 0.91 (0.53-1.56) | 0.72 |
| T1 P1 Control | 333 (24.96) | 284 (86.36) | 0.92 (0.54-1.56) | 0.76 | 1.11 (0.65-1.90) | 0.70 |
| T1 P1 Intervention | 174 (13.04) | 145 (82.34) | 0.68 (0.39-1.19) | 0.17 | 0.68 (0.36-1.28) | 0.23 |
| T1 P2 Control | 28 (2.10) | 21 (74.38) | 0.42 (0.16-1.12) | 0.08 | 0.30 (0.11-0.85) | 0.02 |
| T1 P2 Intervention | 128 (9.60) | 113 (87.46) | 1.01 (0.51-2.01) | 0.97 | 1.38 (0.61-3.11) | 0.44 |
| Missing data | 11 (0.82) | 11 (100.00) | .. | .. | .. | .. |
| **Encounter with a health mediator** | | | | | | |
| Not reported | 1243 (93.18) | 1063 (85.73) | 1 (ref) | .. | 1 (ref) | .. |
| Reported | 60 (4.50) | 52 (88.91) | 1.33 (0.60-2.99) | 0.48 | 1.40 (0.60-3.31) | 0.44 |
| Missing data | 31 (2.32) | 25 (69.05) | .. | .. | .. | .. |
| **Primary care physician** | | | | | | |
| Without | 63 (4.72) | 39 (63.50) | 1 (ref) | .. | 1 (ref) | .. |
| With | 1269 (95.13) | 1100 (86.68) | 3.74 (1.85-7.57) | 0.0003 | 4.46 (2.24-8.89) | <0.0001 |
| Missing data | 2 (0.15) | 1 (50.12) | .. | .. | .. | .. |
| **Age** | | | | | | |
| 50-54 | 369 (27.66) | 291 (80.77) | 1 (ref) | .. | 1 (ref) | .. |
| 55-59 | 265 (19.87) | 233 (89.29) | 1.99 (1.13-3.47) | 0.02 | 1.82 (0.99-3.36) | 0.05 |
| 60-64 | 253 (18.97) | 219 (85.23) | 1.37 (0.79-2.40) | 0.27 | 1.13 (0.63-2.04) | 0.68 |
| 65-69 | 210 (15.74) | 192 (89.99) | 2.14 (1.12-4.07) | 0.02 | 1.55 (0.80-3.00) | 0.19 |
| 70-74 | 237 (17.77) | 205 (88.08) | 1.76 (0.97-3.18) | 0.06 | 1.45 (0.81-2.61) | 0.21 |
| **Socioeconomic profile** | | | | | | |
| NS renter | 644 (48.28) | 573 (89.55) | 1 (ref) | .. | 1 (ref) | .. |
| NS homeowner | 202 (15.14) | 183 (89.11) | 0.95 (0.50-1.82) | 0.89 | 0.90 (0.44-1.83) | 0.77 |
| Non-NS renter | 478 (35.83) | 376 (77.31) | 0.40 (0.26-0.61) | 0.0000 | 0.38 (0.24-0.60) | <0.0001 |
| Missing data[1] | 10 (0.75) | 8 (90.63) | .. | .. | .. | .. |
| **Arrondissement** | | | | | | |
| 1st | 140 (10.49) | 122 (88.87) | 1 (ref) | .. | 1 (ref) | .. |
| 2nd | 172 (12.89) | 149 (87.39) | 0.87 (0.36-2.11) | 0.75 | 1.10 (0.42-2.89) | 0.85 |
| 3rd | 363 (27.21) | 296 (84.14) | 0.66 (0.30-1.46) | 0.31 | 1.07 (0.44-2.60) | 0.87 |
| 13th | 202 (15.14) | 182 (88.41) | 0.96 (0.40-2.26) | 0.92 | 1.20 (0.47-3.06) | 0.70 |
| 14th | 112 (8.40) | 94 (75.16) | 0.38 (0.15-0.94) | 0.04 | 0.44 (0.16-1.23) | 0.12 |
| 15th | 192 (14.39) | 162 (81.61) | 0.56 (0.24-1.27) | 0.16 | 0.71 (0.28-1.80) | 0.48 |
| 16th | 153 (11.47) | 135 (89.30) | 1.05 (0.43-2.52) | 0.92 | 1.09 (0.42-2.85) | 0.86 |

[1], Native speaker student considered as missing data because of to low frequence

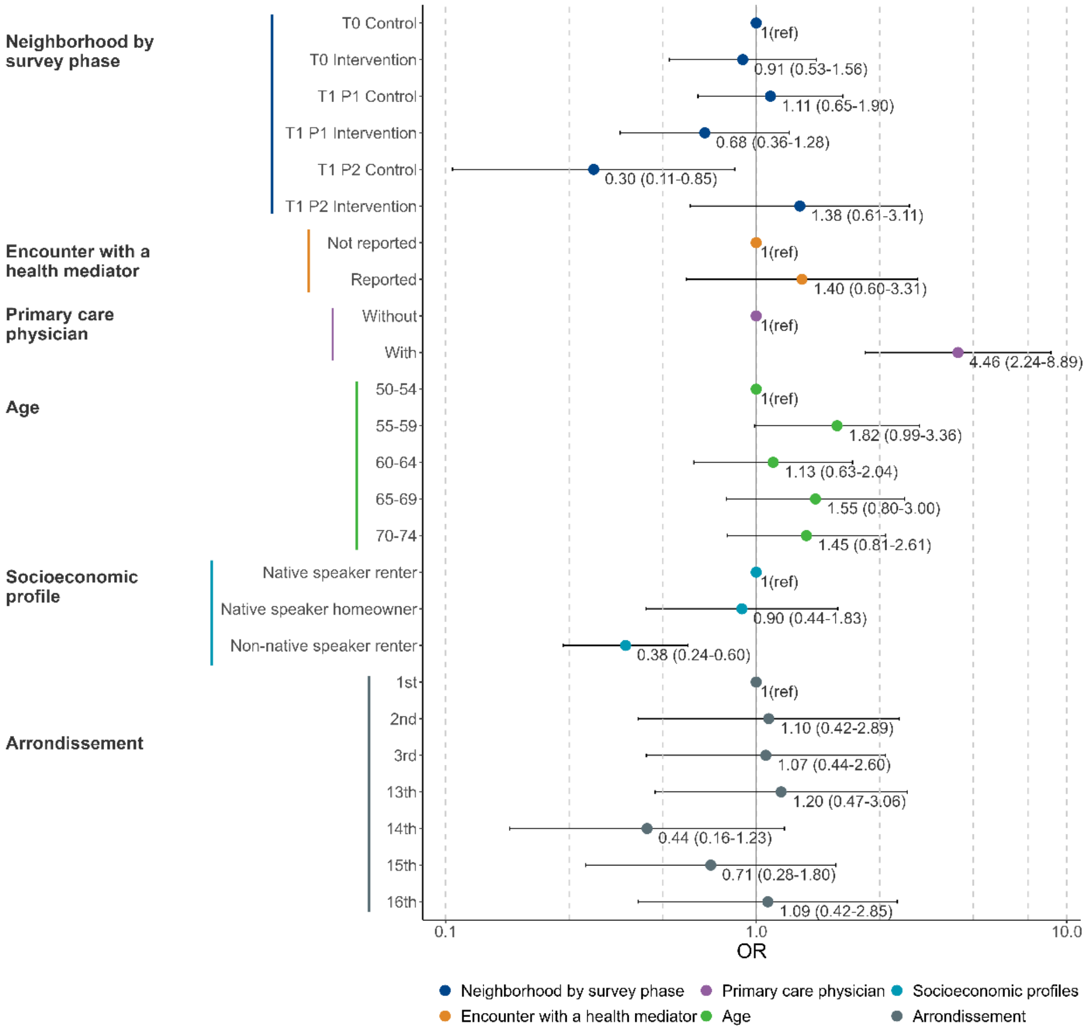

Supplementary figure 13. Factors associated with reception of breast cancer screening invitation, in the multivariate analysis (n=1284).

#### 6.3. Colorectal cancer screening invitation

Supplementary table 4. Factors associated with reception of colorectal cancer screening invitation.

| | n (%) | N receipt invitation (weighted %) | Univariate analysis Odds ratio (95% CI) | Univariate analysis p value | Multivariate analysis Odds ratio (95% CI) | Multivariate analysis p value |
|---|---|---|---|---|---|---|
| **Colorectal cancer screening invitation** (man and women aged 50–74 years; n [receipt invitation] = 1488 (WP=65.15%); n [without missing outcome] = 2034; n [multivariate analysis] = 1963) | | | | | | |
| **Neighborhoods by survey phase** | | | | | | |
| T0 Control | 587 (28.86) | 470 (75.85) | 1 (ref) | .. | 1 (ref) | .. |
| T0 Intervention | 500 (24.58) | 363 (73.41) | 0.88 (0.62-1.24) | 0.46 | 0.85 (0.59-1.23) | 0.39 |
| T1 P1 Control | 453 (22.27) | 329 (72.12) | 0.82 (0.56-1.22) | 0.33 | 0.86 (0.57-1.29) | 0.46 |
| T1 P1 Intervention | 271 (13.32) | 175 (66.56) | 0.63 (0.43-0.94) | 0.02 | 0.57 (0.36-0.92) | 0.02 |
| T1 P2 Control | 45 (2.21) | 25 (56.95) | 0.42 (0.20-0.89) | 0.02 | 0.35 (0.16-0.80) | 0.01 |
| T1 P2 Intervention | 162 (7.96) | 112 (66.90) | 0.64 (0.41-1.01) | 0.06 | 0.61 (0.37-1.00) | 0.05 |
| Missing data | 16 (0.79) | 14 (79.44) | .. | .. | .. | .. |
| **Encounter with a health mediator** | | | | | | |
| Not reported | 1913 (94.05) | 1405 (72.71) | 1 (ref) | .. | 1 (ref) | .. |
| Reported | 87 (4.28) | 64 (64.76) | 0.69 (0.32-1.47) | 0.34 | 1.00 (0.42-2.38) | 1 |
| Missing data | 34 (1.67) | 19 (47.25) | .. | .. | .. | .. |
| **Primary care physician** | | | | | | |
| Without | 119 (5.85) | 58 (48.36) | 1 (ref) | .. | 1 (ref) | .. |
| With | 1913 (94.05) | 1429 (73.73) | 3.00 (1.79-5.00) | <0.0001 | 3.29 (1.85-5.85) | <0.0001 |
| Missing data | 2 (0.10) | 1 (50.12) | .. | .. | .. | .. |
| **Gender** | | | | | | |
| Man | 758 (37.27) | 579 (74.17) | 1 (ref) | .. | 1 (ref) | .. |
| Woman | 1276 (62.73) | 909 (69.72) | 0.80 (0.60-1.08) | 0.14 | 0.69 (0.50-0.94) | 0.02 |
| **Age** | | | | | | |
| 50-54 | 522 (25.66) | 310 (61.28) | 1 (ref) | .. | 1 (ref) | .. |
| 55-59 | 391 (19.22) | 297 (79.40) | 2.44 (1.62-3.67) | <0.0001 | 2.05 (1.31-3.20) | 0.0016 |
| 60-64 | 393 (19.32) | 295 (73.25) | 1.73 (1.15-2.61) | 0.0087 | 1.49 (0.98-2.26) | 0.06 |
| 65-69 | 336 (16.52) | 270 (77.87) | 2.22 (1.41-3.50) | 0.0005 | 1.73 (1.06-2.81) | 0.03 |
| 70-74 | 392 (19.27) | 316 (81.03) | 2.70 (1.78-4.08) | <0.0001 | 2.15 (1.40-3.29) | 0.0004 |
| **Socioeconomic profile** | | | | | | |
| NS renter | 974 (47.89) | 772 (77.78) | 1 (ref) | .. | 1 (ref) | .. |
| NS homeowner | 305 (15.00) | 252 (85.21) | 1.65 (1.05-2.57) | 0.03 | 1.52 (0.94-2.46) | 0.09 |
| Non-NS renter | 733 (36.04) | 451 (57.75) | 0.39 (0.28-0.54) | <0.0001 | 0.39 (0.28-0.55) | <0.0001 |
| Missing data[1] | 22 (1.08) | 13 (61.05) | .. | .. | .. | .. |
| **Arrondissement** | | | | | | |
| 1st | 248 (12.19) | 182 (75.93) | 1 (ref) | .. | 1 (ref) | .. |
| 2nd | 255 (12.54) | 192 (75.73) | 0.99 (0.57-1.72) | 0.97 | 1.07 (0.56-2.04) | 0.84 |
| 3rd | 544 (26.75) | 350 (66.07) | 0.62 (0.39-0.99) | 0.04 | 0.78 (0.46-1.32) | 0.35 |
| 13th | 302 (14.85) | 233 (75.72) | 0.99 (0.60-1.64) | 0.96 | 1.29 (0.73-2.28) | 0.38 |
| 14th | 160 (7.87) | 121 (71.75) | 0.80 (0.43-1.51) | 0.50 | 0.99 (0.45-2.15) | 0.97 |
| 15th | 292 (14.36) | 226 (74.37) | 0.92 (0.56-1.52) | 0.74 | 1.12 (0.64-1.95) | 0.69 |
| 16th | 233 (11.46) | 184 (81.28) | 1.38 (0.81-2.33) | 0.23 | 1.23 (0.69-2.20) | 0.48 |

[1], Native speaker student considered as missing data because of to low frequence

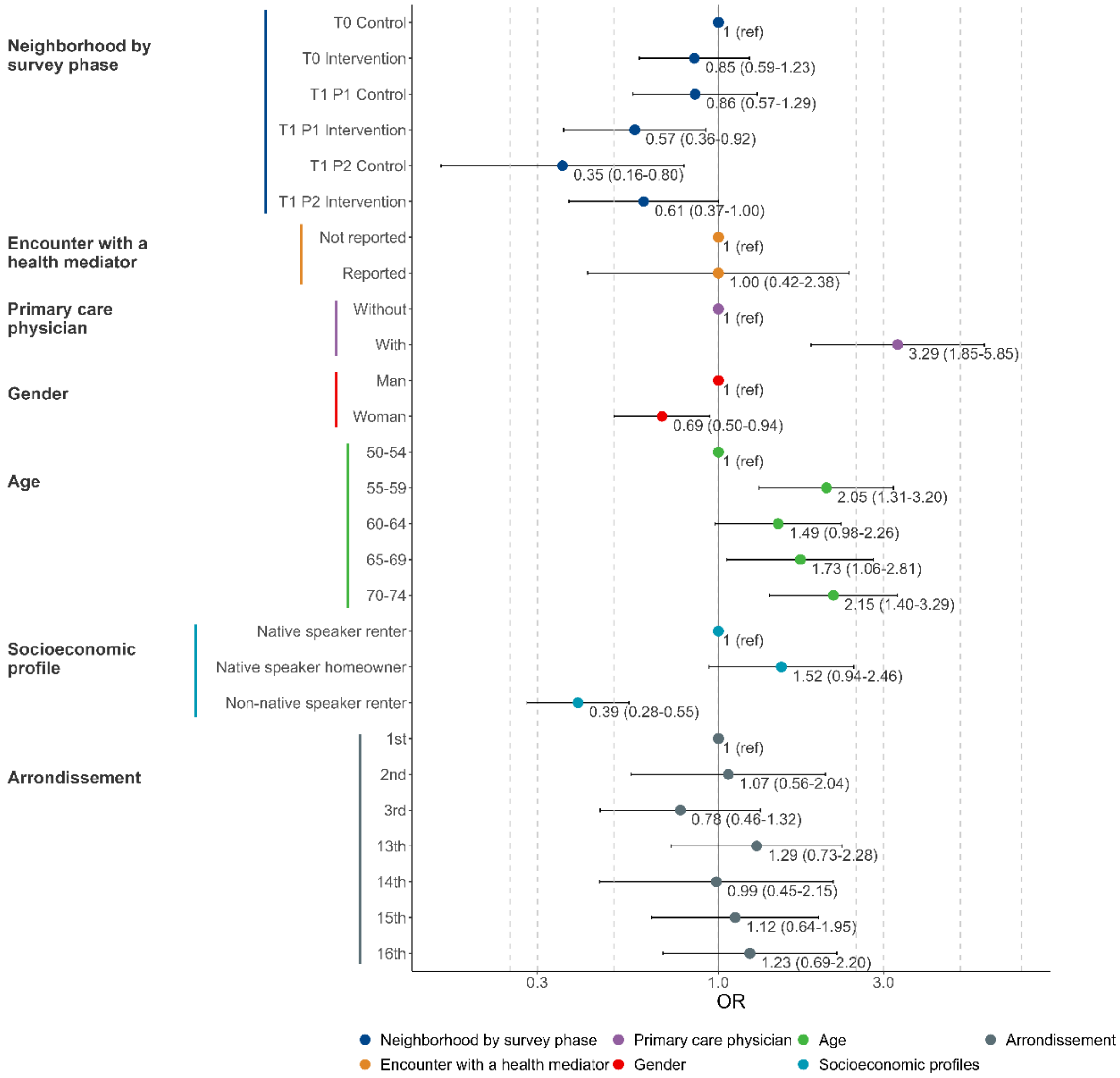

Supplementary figure 14. Factors associated with colorectal cancer screening invitation reception in multivariate analysis (n=1963).

#### 6.4. Cervical cancer screening invitation

Supplementary table 5. Factors associated with reception of cervical cancer screening invitation.

| | n (%) | n receipt invitation (weighted %) | Univariate analysis Odds ratio (95% CI) | p value | Multivariate analysis Odds ratio (95% CI) | p value |
|---|---|---|---|---|---|---|
| **Cervical cancer screening invitation** (women aged 25–64 years; n [receipt invitation] = 1088 (WP=53.30%); n [without missing outcome] = 1775; n; n [multivariate analysis] = 1731) | | | | | | |
| **Neighborhoods by survey phase** | | | | | | |
| T0 Control | 487 (27.44) | 300 (59.27) | 1 (ref) | .. | 1 (ref) | .. |
| T0 Intervention | 464 (26.14) | 239 (51.13) | 0.72 (0.53-0.97) | 0.03 | 0.71 (0.51-0.98) | 0.04 |
| T1 P1 Control | 393 (22.14) | 258 (66.00) | 1.33 (0.95-1.87) | 0.09 | 1.37 (0.96-1.95) | 0.08 |
| T1 P1 Intervention | 242 (13.63) | 158 (64.95) | 1.27 (0.89-1.82) | 0.18 | 1.07 (0.72-1.60) | 0.74 |
| T1 P2 Control | 36 (2.03) | 17 (54.60) | 0.83 (0.40-1.73) | 0.61 | 0.68 (0.32-1.44) | 0.31 |
| T1 P2 Intervention | 135 (7.61) | 103 (76.32) | 2.21 (1.37-3.57) | 0.0011 | 1.79 (1.04-3.07) | 0.04 |
| Missing data | 18 (1.01) | 13 (59.26) | .. | .. | .. | .. |
| **Encounter with a health mediator** | | | | | | |
| Not reported | 1668 (93.97) | 1008 (60.93) | 1 (ref) | .. | 1 (ref) | .. |
| Reported | 84 (4.73) | 65 (78.80) | 2.38 (1.28-4.43) | 0.0061 | 2.00 (1.03-3.88) | 0.04 |
| Missing data | 23 (1.30) | 15 (49.51) | .. | .. | .. | .. |
| **Primary care physician** | | | | | | |
| Without | 146 (8.23) | 57 (40.21) | 1 (ref) | .. | 1 (ref) | .. |
| With | 1627 (91.66) | 1030 (63.63) | 2.60 (1.68-4.02) | <0.0001 | 2.57 (1.59-4.14) | 0.0001 |
| Missing data | 2 (0.11) | 1 (50.12) | .. | .. | .. | .. |
| **Age** | | | | | | |
| 25-29 | 237 (13.35) | 131 (53.01) | 1 (ref) | .. | 1 (ref) | .. |
| 30-34 | 197 (11.10) | 126 (64.45) | 1.61 (0.97-2.66) | 0.07 | 1.80 (1.03-3.14) | 0.04 |
| 35-39 | 229 (12.90) | 143 (64.11) | 1.58 (0.95-2.63) | 0.08 | 1.84 (1.05-3.24) | 0.03 |
| 40-44 | 167 (9.41) | 102 (61.16) | 1.40 (0.83-2.34) | 0.21 | 1.61 (0.90-2.91) | 0.11 |
| 45-49 | 186 (10.48) | 103 (57.29) | 1.19 (0.70-2.01) | 0.52 | 1.26 (0.69-2.27) | 0.45 |
| 50-54 | 315 (17.75) | 198 (64.96) | 1.64 (1.05-2.56) | 0.03 | 1.88 (1.15-3.07) | 0.01 |
| 55-59 | 230 (12.96) | 150 (66.79) | 1.78 (1.10-2.89) | 0.02 | 1.81 (1.07-3.07) | 0.03 |
| 60-64 | 214 (12.06) | 135 (60.02) | 1.33 (0.82-2.15) | 0.24 | 1.25 (0.73-2.12) | 0.41 |
| **Socioeconomic profile** | | | | | | |
| NS renter | 809 (45.58) | 569 (69.33) | 1 (ref) | .. | 1 (ref) | .. |
| NS homeowner | 179 (10.08) | 112 (60.83) | 0.69 (0.45-1.05) | 0.08 | 0.64 (0.41-1.00) | 0.05 |
| NS student | 87 (4.90) | 45 (55.25) | 0.55 (0.31-0.97) | 0.04 | 0.72 (0.39-1.35) | 0.31 |
| Non-NS renter | 699 (39.38) | 361 (52.41) | 0.49 (0.37-0.65) | <0.0001 | 0.45 (0.33-0.61) | <0.0001 |
| Missing data | 1 (0.06) | 1 (100.00) | .. | .. | .. | .. |
| **Arrondissement** | | | | | | |
| 1st | 199 (11.21) | 125 (61.21) | 1 (ref) | .. | 1 (ref) | .. |
| 2nd | 219 (12.34) | 135 (64.22) | 1.14 (0.67-1.94) | 0.63 | 1.12 (0.61-2.03) | 0.72 |
| 3rd | 515 (29.01) | 275 (57.85) | 0.87 (0.55-1.37) | 0.55 | 0.92 (0.54-1.58) | 0.76 |
| 13th | 259 (14.59) | 178 (67.67) | 1.33 (0.81-2.17) | 0.26 | 1.17 (0.66-2.07) | 0.59 |
| 14th | 173 (9.75) | 121 (65.73) | 1.22 (0.71-2.09) | 0.48 | 1.15 (0.63-2.11) | 0.65 |
| 15th | 213 (12.00) | 140 (67.06) | 1.29 (0.78-2.14) | 0.32 | 1.16 (0.65-2.05) | 0.61 |
| 16th | 197 (11.10) | 114 (60.75) | 0.98 (0.59-1.63) | 0.94 | 0.82 (0.46-1.48) | 0.52 |

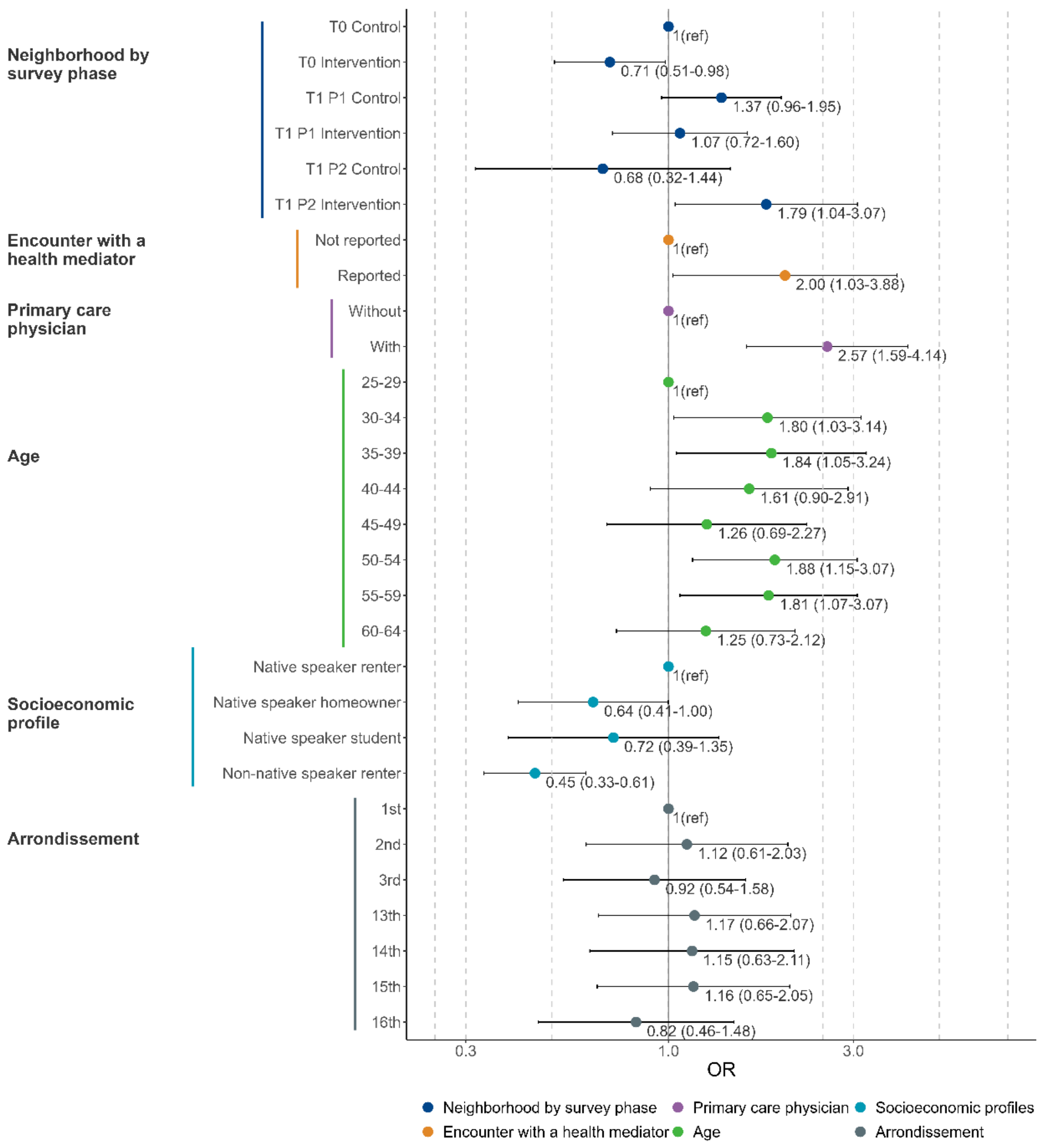

Supplementary figure 15. Factors associated with cervical cancer screening invitation reception in multivariate analysis (n=).

### APPENDIX 7. ENCOUNTER WITH A HEALTH MEDIATOR

Supplementary table 6. Factors associated with individuals reporting an encounter with a health mediator during the T1 survey.

| | n (%) | n with encounter (weighted %) | Univariate analysis Odds ratio (95% CI) | p value | Multivariate analysis Odds ratio (95% CI) | p value |
|---|---|---|---|---|---|---|
| **Encounter with a health mediator** (n [without missing outcome] = 1805; n [multivariable analysis] = 1764) | | | | | | |
| **Neighborhoods by survey phase** | | | | | | |
| T1 P1 Control | 851 (47.15) | 42 (3.80) | 1 (ref) | .. | 1 (ref) | .. |
| T1 P1 Intervention | 571 (31.63) | 96 (16.62) | 5.05 (2.89-8.82) | <0.0001 | 5.43 (3.10-9.51) | <0.0001 |
| T1 P2 Control | 79 (4.38) | 2 (2.26) | 0.59 (0.12-2.93) | 0.5149 | 0.72 (0.15-3.46) | 0.68 |
| T1 P2 Intervention | 265 (14.68) | 14 (3.65) | 0.96 (0.43-2.12) | 0.9160 | 1.31 (0.50-3.40) | 0.58 |
| Missing data | 39 (2.16) | 9 (17.93) | .. | .. | .. | .. |
| **Primary care physician** | | | | | | |
| Without | 131 (7.26) | 7 (3.12) | 1 (ref) | .. | 1 (ref) | .. |
| With | 1672 (92.63) | 155 (6.59) | 2.19 (0.83-5.80) | 0.11 | 2.12 (0.71-6.30) | 0.18 |
| Missing data | 2 (0.11) | 1 (53.29) | .. | .. | .. | .. |
| **Gender** | | | | | | |
| Man | 618 (34.24) | 51 (6.22) | 1 (ref) | .. | 1 (ref) | .. |
| Woman | 1187 (65.76) | 112 (6.31) | 1.02 (0.62-1.66) | 0.95 | 0.96 (0.55-1.67) | 0.87 |
| **Age** | | | | | | |
| 18-24 | 149 (8.25) | 11 (5.81) | 1 (ref) | .. | 1 (ref) | .. |
| 25-29 | 180 (9.97) | 9 (3.10) | 0.52 (0.18-1.49) | 0.22 | 0.60 (0.22-1.63) | 0.32 |
| 30-34 | 129 (7.15) | 8 (3.48) | 0.58 (0.21-1.65) | 0.31 | 1.02 (0.35-2.92) | 0.98 |
| 35-39 | 131 (7.26) | 13 (9.04) | 1.61 (0.48-5.45) | 0.44 | 2.74 (0.75-9.94) | 0.13 |
| 40-44 | 102 (5.65) | 14 (6.64) | 1.15 (0.44-3.02) | 0.77 | 2.19 (0.77-6.24) | 0.14 |
| 45-49 | 115 (6.37) | 12 (8.17) | 1.44 (0.56-3.73) | 0.45 | 2.01 (0.72-5.65) | 0.18 |
| 50-54 | 267 (14.79) | 28 (8.07) | 1.42 (0.58-3.47) | 0.44 | 2.01 (0.70-5.77) | 0.20 |
| 55-59 | 174 (9.64) | 18 (6.22) | 1.08 (0.45-2.59) | 0.87 | 1.95 (0.74-5.17) | 0.18 |
| 60-64 | 217 (12.02) | 22 (7.73) | 1.36 (0.55-3.33) | 0.50 | 2.10 (0.74-5.93) | 0.16 |
| 65-69 | 170 (9.42) | 16 (5.79) | 1.00 (0.40-2.46) | 0.99 | 1.66 (0.62-4.42) | 0.31 |
| 70-74 | 171 (9.47) | 12 (5.58) | 0.96 (0.36-2.53) | 0.93 | 1.52 (0.47-4.96) | 0.48 |
| **Socioeconomic profile** | | | | | | |
| NS renter | 784 (43.43) | 44 (3.68) | 1 (ref) | .. | 1 (ref) | .. |
| NS homeowner | 238 (13.19) | 15 (4.18) | 1.14 (0.54-2.41) | 0.73 | 1.54 (0.69-3.44) | 0.29 |
| NS student | 186 (10.30) | 15 (6.26) | 1.75 (0.85-3.59) | 0.13 | 2.80 (1.25-6.27) | 0.01 |
| Non-NS renter | 596 (33.02) | 88 (10.81) | 3.18 (1.84-5.49) | <0.0001 | 2.84 (1.63-4.96) | 0.0002 |
| Missing data | 1 (0.06) | 1 (100.00) | .. | .. | .. | .. |
| **Arrondissement** | | | | | | |
| 1st | 226 (12.52) | 24 (5.07) | 1 (ref) | .. | 1 (ref) | .. |
| 2nd | 215 (11.91) | 18 (4.95) | 0.98 (0.44-2.15) | 0.95 | 0.98 (0.42-2.30) | 0.96 |
| 3rd | 470 (26.04) | 60 (6.81) | 1.37 (0.69-2.72) | 0.37 | 1.59 (0.76-3.30) | 0.22 |
| 13th | 301 (16.68) | 34 (11.11) | 2.34 (1.20-4.57) | 0.01 | 1.61 (0.75-3.45) | 0.22 |
| 14th | 148 (8.20) | 15 (10.88) | 2.29 (1.01-5.16) | 0.05 | 1.55 (0.62-3.87) | 0.35 |
| 15th | 251 (13.91) | 6 (3.21) | 0.62 (0.22-1.75) | 0.37 | 1.00 (0.29-3.40) | 1 |
| 16th | 194 (10.75) | 6 (2.40) | 0.46 (0.17-1.26) | 0.13 | 0.30 (0.10-0.89) | 0.03 |

### APPENDIX 8. CANCER SCREENING UPTAKE

#### 8.1. Breast cancer screening uptake

Supplementary table 7. Factors associated with up-to-date breast cancer screening.

| | n (%) | n up to date (weighted %) | Univariate analysis Odds ratio (95% CI) | Univariate analysis p value | Multivariate analysis Odds ratio (95% CI) | Multivariate analysis p value |
|---|---|---|---|---|---|---|
| **Breast cancer screening uptake** (women aged 50–74 years, n [without missing outcome] = 1 389, n [multivariate analysis] = 1252) | | | | | | |
| **Neighborhoods by survey phase** | | | | | | |
| T0 Control | 379 (27.29) | 191 (52.82) | 1 (ref) | .. | 1 (ref) | .. |
| T0 Intervention | 321 (23.11) | 181 (57.66) | 1.22 (0.86-1.72) | 0.28 | 1.43 (0.97-2.13) | 0.07 |
| T1 P1 Control | 340 (24.48) | 217 (63.32) | 1.54 (1.06-2.23) | 0.02 | 1.37 (0.90-2.07) | 0.14 |
| T1 P1 Intervention | 182 (13.10) | 96 (54.42) | 1.07 (0.71-1.59) | 0.75 | 0.90 (0.56-1.45) | 0.67 |
| T1 P2 Control | 29 (2.09) | 5 (24.47) | 0.29 (0.10-0.81) | 0.02 | 0.27 (0.10-0.76) | 0.01 |
| T1 P2 Intervention | 126 (9.07) | 41 (29.31) | 0.37 (0.23-0.59) | <0.0001 | 0.28 (0.16-0.49) | <0.0001 |
| Missing data | 12 (0.86) | 7 (70.46) | .. | .. | .. | .. |
| **Encounter with a health mediator** | | | | | | |
| Not reported | 1292 (93.02) | 679 (53.76) | 1 (ref) | .. | 1 (ref) | .. |
| Reported | 61 (4.39) | 42 (73.62) | 2.40 (1.31-4.41) | 0.0048 | 2.26 (1.12-4.54) | 0.02 |
| Missing data | 36 (2.59) | 17 (58.01) | .. | .. | .. | .. |
| **Primary care physician** | | | | | | |
| Without | 65 (4.68) | 17 (29.75) | 1 (ref) | .. | 1 (ref) | .. |
| With | 1322 (95.18) | 721 (55.96) | 3.00 (1.41-6.40) | 0.0045 | 3.90 (1.76-8.65) | 0.0008 |
| Missing data | 2 (0.14) | 0 (0.00) | .. | .. | .. | .. |
| **Screening invitation** | | | | | | |
| Not received | 187 (13.46) | 74 (38.81) | 1 (ref) | .. | 1 (ref) | .. |
| Received | 1115 (80.27) | 646 (58.83) | 2.25 (1.50-3.39) | 0.0001 | 2.52 (1.67-3.78) | <0.0001 |
| Missing data | 87 (6.26) | 18 (20.18) | .. | .. | .. | .. |
| **Age** | | | | | | |
| 50-54 | 377 (27.14) | 226 (63.20) | 1 (ref) | .. | 1 (ref) | .. |
| 55-59 | 277 (19.94) | 158 (53.65) | 0.67 (0.44-1.03) | 0.07 | 0.51 (0.31-0.83) | 0.0064 |
| 60-64 | 264 (19.01) | 129 (49.65) | 0.57 (0.38-0.86) | 0.0068 | 0.50 (0.32-0.78) | 0.0022 |
| 65-69 | 221 (15.91) | 122 (55.59) | 0.73 (0.48-1.11) | 0.14 | 0.62 (0.38-1.01) | 0.05 |
| 70-74 | 250 (18.00) | 103 (39.36) | 0.38 (0.25-0.57) | <0.0001 | 0.30 (0.19-0.49) | <0.0001 |
| **Socioeconomic profile** | | | | | | |
| NS renter | 659 (47.44) | 361 (53.30) | 1 (ref) | .. | 1 (ref) | .. |
| NS homeowner | 205 (14.76) | 116 (57.02) | 1.16 (0.78-1.74) | 0.46 | 1.17 (0.74-1.84) | 0.50 |
| Non-NS renter | 514 (37.01) | 258 (54.72) | 1.06 (0.78-1.44) | 0.72 | 1.02 (0.72-1.46) | 0.90 |
| Missing data[1] | 11 (0.79) | 3 (57.23) | .. | .. | .. | .. |
| **Arrondissement** | | | | | | |
| 1st | 147 (10.58) | 79 (63.47) | 1 (ref) | .. | 1 (ref) | .. |
| 2nd | 182 (13.10) | 83 (43.23) | 0.44 (0.24-0.80) | 0.0068 | 0.58 (0.29-1.18) | 0.13 |
| 3rd | 369 (26.57) | 201 (55.33) | 0.71 (0.42-1.21) | 0.21 | 0.67 (0.36-1.24) | 0.20 |
| 13th | 214 (15.41) | 117 (55.65) | 0.72 (0.42-1.25) | 0.24 | 0.77 (0.41-1.44) | 0.41 |
| 14th | 117 (8.42) | 58 (49.08) | 0.55 (0.30-1.03) | 0.06 | 0.54 (0.27-1.10) | 0.09 |
| 15th | 211 (15.19) | 101 (49.18) | 0.56 (0.32-0.96) | 0.04 | 0.78 (0.41-1.47) | 0.44 |
| 16th | 149 (10.73) | 99 (67.31) | 1.19 (0.66-2.13) | 0.57 | 1.10 (0.57-2.14) | 0.77 |

[1], Native speaker student considered as missing data because of to low frequence

#### 8.2. Colorectal cancer screening uptake

Supplementary table 8. Factors associated with up-to-date colorectal cancer screening.

| | n (%) | n up to date (weighted %) | Univariate analysis Odds ratio (95% CI) | p value | Multivariate analysis Odds ratio (95% CI) | p value |
|---|---|---|---|---|---|---|
| **Colorectal cancer screening uptake** (man and women aged 50–74 years, n [without missing outcome] = 2206, n [multivariate analysis] = 1925) | | | | | | |
| **Neighborhoods by survey phase** | | | | | | |
| T0 Control | 632 (28.65) | 209 (29.33) | 1 (ref) | .. | 1 (ref) | .. |
| T0 Intervention | 545 (24.71) | 170 (29.34) | 1.00 (0.74-1.36) | 1 | 1.03 (0.72-1.47) | 0.89 |
| T1 P1 Control | 486 (22.03) | 163 (33.93) | 1.24 (0.88-1.75) | 0.23 | 1.14 (0.78-1.67) | 0.50 |
| T1 P1 Intervention | 304 (13.78) | 84 (31.22) | 1.09 (0.76-1.58) | 0.63 | 0.91 (0.59-1.39) | 0.65 |
| T1 P2 Control | 45 (2.04) | 5 (11.98) | 0.33 (0.12-0.92) | 0.03 | 0.39 (0.12-1.21) | 0.10 |
| T1 P2 Intervention | 172 (7.80) | 39 (21.61) | 0.66 (0.42-1.05) | 0.08 | 0.69 (0.40-1.20) | 0.19 |
| Missing data | 22 (1.00) | 8 (42.49) | .. | .. | .. | .. |
| **Encounter with a health mediator** | | | | | | |
| Not reported | 2062 (93.47) | 626 (29.55) | 1 (ref) | .. | 1 (ref) | .. |
| Reported | 93 (4.22) | 43 (50.01) | 2.38 (1.29-4.41) | 0.0056 | 2.75 (1.31-5.77) | 0.0074 |
| Missing data | 51 (2.31) | 9 (16.96) | .. | .. | .. | .. |
| **Primary care physician** | | | | | | |
| Without | 131 (5.94) | 10 (10.34) | 1 (ref) | .. | 1 (ref) | .. |
| With | 2073 (93.97) | 667 (31.35) | 3.96 (1.72-9.10) | 0.0012 | 3.02 (1.26-7.26) | 0.01 |
| Missing data | 2 (0.09) | 1 (50.12) | .. | .. | .. | .. |
| **Screening invitation** | | | | | | |
| Not received | 529 (23.98) | 63 (11.92) | 1 (ref) | .. | 1 (ref) | .. |
| Received | 1465 (66.41) | 595 (38.88) | 4.70 (3.18-6.93) | <0.0001 | 4.25 (2.74-6.58) | <0.0001 |
| Missing data | 212 (9.61) | 20 (13.44) | .. | .. | .. | .. |
| **Gender** | | | | | | |
| Man | 823 (37.31) | 270 (33.69) | 1 (ref) | .. | 1 (ref) | .. |
| Woman | 1383 (62.69) | 408 (26.22) | 0.70 (0.54-0.91) | 0.0075 | 0.69 (0.52-0.92) | 0.01 |
| **Age** | | | | | | |
| 50-54 | 571 (25.88) | 156 (25.85) | 1 (ref) | .. | 1 (ref) | .. |
| 55-59 | 424 (19.22) | 130 (29.17) | 1.18 (0.79-1.77) | 0.42 | 0.92 (0.58-1.46) | 0.73 |
| 60-64 | 421 (19.08) | 141 (35.20) | 1.56 (1.05-2.32) | 0.03 | 1.30 (0.84-2.01) | 0.23 |
| 65-69 | 365 (16.55) | 125 (38.56) | 1.80 (1.17-2.76) | 0.0070 | 1.54 (0.94-2.52) | 0.09 |
| 70-74 | 425 (19.27) | 126 (26.89) | 1.06 (0.71-1.56) | 0.79 | 0.85 (0.56-1.30) | 0.46 |
| **Socioeconomic profile** | | | | | | |
| NS renter | 1043 (47.28) | 345 (30.79) | 1 (ref) | .. | 1 (ref) | .. |
| NS homeowner | 311 (14.10) | 105 (36.83) | 1.31 (0.91-1.88) | 0.14 | 1.40 (0.92-2.14) | 0.12 |
| Non-NS renter | 828 (37.53) | 223 (25.85) | 0.78 (0.57-1.08) | 0.13 | 0.96 (0.66-1.39) | 0.81 |
| Missing data[1] | 24 (1.09) | 5 (20.71) | .. | .. | .. | .. |
| **Arrondissement** | | | | | | |
| 1st | 263 (11.92) | 69 (29.01) | 1 (ref) | .. | 1 (ref) | .. |
| 2nd | 269 (12.19) | 63 (25.81) | 0.85 (0.45-1.61) | 0.62 | 0.92 (0.45-1.87) | 0.81 |
| 3rd | 577 (26.16) | 149 (26.82) | 0.90 (0.56-1.44) | 0.65 | 1.02 (0.59-1.76) | 0.95 |
| 13th | 344 (15.59) | 139 (38.06) | 1.50 (0.93-2.42) | 0.09 | 1.68 (0.94-2.97) | 0.08 |
| 14th | 183 (8.30) | 63 (36.82) | 1.43 (0.82-2.48) | 0.21 | 1.69 (0.88-3.24) | 0.12 |
| 15th | 332 (15.05) | 120 (35.82) | 1.37 (0.84-2.22) | 0.21 | 1.68 (0.96-2.93) | 0.09 |
| 16th | 238 (10.79) | 75 (34.30) | 1.28 (0.77-2.12) | 0.34 | 1.14 (0.65-2.00) | 0.65 |

[1], Native speaker student considered as missing data because of to low frequence

#### 8.3. Cervical cancer screening uptake

Supplementary table 9. Factors associated with up-to-date cervical cancer screening.

| | n (%) | n up to date (weighted %) | Univariate analysis Odds ratio (95% CI) | p value | Multivariate analysis Odds ratio (95% CI) | p value |
|---|---|---|---|---|---|---|
| **Cervical cancer screening uptake** (women aged 25–64 years, n [without missing outcome] = 2001, n [multivariate analysis] = 1666) | | | | | | |
| **Neighborhoods by survey phase** | | | | | | |
| T0 Control | 586 (29.29) | 404 (70.97) | 1 (ref) | .. | 1 (ref) | .. |
| T0 Intervention | 538 (26.89) | 394 (72.97) | 1.10 (0.82-1.49) | 0.52 | 1.10 (0.76-1.60) | 0.62 |
| T1 P1 Control | 423 (21.14) | 315 (75.02) | 1.23 (0.87-1.73) | 0.24 | 0.97 (0.64-1.46) | 0.87 |
| T1 P1 Intervention | 253 (12.64) | 180 (71.36) | 1.02 (0.71-1.46) | 0.92 | 0.85 (0.54-1.34) | 0.49 |
| T1 P2 Control | 35 (1.75) | 18 (55.07) | 0.50 (0.24-1.06) | 0.07 | 0.44 (0.19-1.00) | 0.05 |
| T1 P2 Intervention | 145 (7.25) | 82 (55.86) | 0.52 (0.34-0.78) | 0.0018 | 0.46 (0.27-0.79) | 0.0050 |
| Missing data | 21 (1.05) | 15 (83.21) | .. | .. | .. | .. |
| **Encounter with a health mediator** | | | | | | |
| Not reported | 1884 (94.15) | 1319 (71.00) | 1 (ref) | .. | 1 (ref) | .. |
| Reported | 87 (4.35) | 72 (83.46) | 2.06 (1.04-4.08) | 0.04 | 1.79 (0.82-3.89) | 0.14 |
| Missing data | 30 (1.50) | 17 (67.75) | .. | .. | .. | .. |
| **Primary care physician** | | | | | | |
| Without | 169 (8.45) | 96 (63.46) | 1 (ref) | .. | 1 (ref) | .. |
| With | 1830 (91.45) | 1311 (72.23) | 1.50 (1.00-2.25) | 0.05 | 1.39 (0.84-2.30) | 0.19 |
| Missing data | 2 (0.10) | 1 (50.12) | .. | .. | .. | .. |
| **Screening invitation** | | | | | | |
| Not received | 654 (32.68) | 414 (64.04) | 1 (ref) | .. | 1 (ref) | .. |
| Received | 1055 (52.72) | 836 (79.05) | 2.12 (1.59-2.83) | <0.0001 | 2.20 (1.61-3.00) | <0.0001 |
| Missing data | 292 (14.59) | 158 (56.46) | .. | .. | .. | .. |
| **Age** | | | | | | |
| 25-29 | 264 (13.19) | 140 (55.00) | 1 (ref) | .. | 1 (ref) | .. |
| 30-34 | 227 (11.34) | 181 (81.39) | 3.58 (2.08-6.16) | <0.0001 | 2.78 (1.43-5.41) | 0.0026 |
| 35-39 | 254 (12.69) | 201 (80.79) | 3.44 (2.03-5.84) | <0.0001 | 2.29 (1.25-4.22) | 0.0075 |
| 40-44 | 180 (9.00) | 137 (71.59) | 2.06 (1.20-3.53) | 0.0084 | 1.33 (0.69-2.57) | 0.39 |
| 45-49 | 210 (10.49) | 162 (79.37) | 3.15 (1.86-5.32) | <0.0001 | 1.95 (1.05-3.62) | 0.04 |
| 50-54 | 356 (17.79) | 259 (73.67) | 2.29 (1.48-3.55) | 0.0002 | 1.59 (0.93-2.71) | 0.09 |
| 55-59 | 256 (12.79) | 190 (72.06) | 2.11 (1.25-3.55) | 0.0050 | 1.44 (0.76-2.71) | 0.26 |
| 60-64 | 254 (12.69) | 138 (56.87) | 1.08 (0.69-1.68) | 0.74 | 0.72 (0.42-1.22) | 0.22 |
| **Socioeconomic profile** | | | | | | |
| NS renter | 902 (45.08) | 660 (72.86) | 1 (ref) | .. | 1 (ref) | .. |
| NS homeowner | 196 (9.80) | 160 (84.61) | 2.05 (1.26-3.34) | 0.0040 | 1.94 (1.11-3.39) | 0.02 |
| NS student | 100 (5.00) | 34 (30.83) | 0.17 (0.09-0.29) | <0.0001 | 0.21 (0.11-0.40) | <0.0001 |
| Non-NS renter | 802 (40.08) | 553 (70.66) | 0.90 (0.68-1.19) | 0.45 | 1.01 (0.72-1.42) | 0.95 |
| Missing data | 1 (0.05) | 1 (100.00) | .. | .. | .. | .. |
| **Arrondissement** | | | | | | |
| 1st | 219 (10.94) | 159 (77.59) | 1 (ref) | .. | 1 (ref) | .. |
| 2nd | 231 (11.54) | 173 (75.66) | 0.90 (0.52-1.54) | 0.70 | 0.77 (0.40-1.46) | 0.42 |
| 3rd | 555 (27.74) | 394 (69.32) | 0.65 (0.42-1.03) | 0.06 | 0.57 (0.32-1.00) | 0.05 |
| 13th | 301 (15.04) | 191 (64.42) | 0.52 (0.33-0.83) | 0.0066 | 0.56 (0.31-1.01) | 0.05 |
| 14th | 178 (8.90) | 129 (73.58) | 0.80 (0.47-1.38) | 0.43 | 0.63 (0.34-1.18) | 0.15 |
| 15th | 291 (14.54) | 180 (64.43) | 0.52 (0.33-0.83) | 0.0066 | 0.57 (0.31-1.03) | 0.06 |
| 16th | 226 (11.29) | 182 (79.47) | 1.12 (0.66-1.90) | 0.68 | 0.92 (0.48-1.75) | 0.79 |

### APPENDIX 9 CANCER SCREENING AWARENESS

#### 9.1. Colorectal cancer screening awareness

Supplementary table 10. Factors associated with awareness about colorectal cancer screening.

| | n (%) | n aware (weighted %) | Univariate analysis Odds ratio (95% CI) | p value | Multivariate analysis Odds ratio (95% CI) | p value |
|---|---|---|---|---|---|---|
| **Colorectal cancer screening awareness** (man and women aged 50–74 years, n [without missing outcome] = 2273, n [multivariate analysis] =1959) | | | | | | |
| **Neighborhoods by survey phase** | | | | | | |
| T0 Control | 655 (28.82) | 562 (83.76) | 1 (ref) | .. | 1 (ref) | .. |
| T0 Intervention | 572 (25.16) | 456 (80.58) | 0.80 (0.57-1.14) | 0.23 | 1.01 (0.58-1.76) | 0.98 |
| T1 P1 Control | 496 (21.82) | 445 (87.28) | 1.33 (0.79-2.23) | 0.28 | 1.16 (0.62-2.14) | 0.65 |
| T1 P1 Intervention | 308 (13.55) | 227 (77.57) | 0.67 (0.45-1.01) | 0.05 | 0.57 (0.30-1.07) | 0.08 |
| T1 P2 Control | 46 (2.02) | 34 (74.21) | 0.56 (0.25-1.27) | 0.16 | 0.56 (0.23-1.34) | 0.19 |
| T1 P2 Intervention | 174 (7.66) | 141 (80.38) | 0.79 (0.48-1.31) | 0.36 | 0.88 (0.42-1.84) | 0.73 |
| Missing data | 22 (0.97) | 20 (92.15) | .. | .. | .. | .. |
| **Encounter with a health mediator** | | | | | | |
| Not reported | 2125 (93.49) | 1753 (82.81) | 1 (ref) | .. | 1 (ref) | .. |
| Reported | 96 (4.22) | 86 (93.43) | 2.95 (1.41-6.18) | 0.0042 | 8.07 (2.10-30.96) | 0.0023 |
| Missing data | 52 (2.29) | 46 (88.28) | .. | .. | .. | .. |
| **Primary care physician** | | | | | | |
| Without | 137 (6.03) | 91 (73.67) | 1 (ref) | .. | 1 (ref) | .. |
| With | 2133 (93.84) | 1792 (84.08) | 1.89 (1.15-3.09) | 0.01 | 0.89 (0.41-1.94) | 0.77 |
| Missing data | 3 (0.13) | 2 (88.65) | .. | .. | .. | .. |
| **Screening invitation** | | | | | | |
| Not received | 544 (23.93) | 314 (56.49) | 1 (ref) | .. | 1 (ref) | .. |
| Received | 1486 (65.38) | 1438 (96.71) | 22.67 (14.25-36.07) | <0.0001 | 20.77 (12.57-34.34) | <0.0001 |
| Missing data | 243 (10.69) | 133 (62.19) | .. | .. | .. | .. |
| **Gender** | | | | | | |
| Man | 843 (37.09) | 693 (82.77) | 1 (ref) | .. | 1 (ref) | .. |
| Woman | 1430 (62.91) | 1192 (83.93) | 1.09 (0.78-1.52) | 0.62 | 1.62 (1.02-2.57) | 0.04 |
| **Age** | | | | | | |
| 50-54 | 592 (26.04) | 447 (77.46) | 1 (ref) | .. | 1 (ref) | .. |
| 55-59 | 437 (19.23) | 363 (85.81) | 1.76 (1.12-2.77) | 0.01 | 0.88 (0.48-1.61) | 0.67 |
| 60-64 | 432 (19.01) | 365 (85.42) | 1.71 (1.08-2.68) | 0.02 | 1.47 (0.81-2.68) | 0.21 |
| 65-69 | 371 (16.32) | 328 (90.20) | 2.68 (1.60-4.48) | 0.0002 | 1.78 (0.88-3.61) | 0.11 |
| 70-74 | 441 (19.40) | 382 (85.91) | 1.78 (1.12-2.82) | 0.01 | 0.91 (0.49-1.70) | 0.76 |
| **Socioeconomic profile** | | | | | | |
| NS renter | 1074 (47.25) | 949 (89.14) | 1 (ref) | .. | 1 (ref) | .. |
| NS homeowner | 314 (13.81) | 297 (93.93) | 1.88 (0.96-3.70) | 0.07 | 1.51 (0.66-3.45) | 0.33 |
| Non-NS renter | 860 (37.84) | 617 (70.81) | 0.30 (0.21-0.42) | <0.0001 | 0.34 (0.21-0.54) | <0.0001 |
| Missing data[1] | 25 (1.10) | 22 (91.18) | .. | .. | .. | .. |
| **Arrondissement** | | | | | | |
| 1st | 267 (11.75) | 221 (86.21) | 1 (ref) | .. | 1 (ref) | .. |
| 2nd | 274 (12.05) | 237 (85.45) | 0.94 (0.49-1.82) | 0.85 | 0.86 (0.35-2.16) | 0.76 |
| 3rd | 597 (26.26) | 471 (80.50) | 0.66 (0.39-1.13) | 0.13 | 0.70 (0.33-1.50) | 0.36 |
| 13th | 356 (15.66) | 296 (83.52) | 0.81 (0.47-1.40) | 0.45 | 1.15 (0.50-2.65) | 0.74 |
| 14th | 185 (8.14) | 151 (82.07) | 0.73 (0.36-1.48) | 0.38 | 0.90 (0.33-2.43) | 0.83 |
| 15th | 349 (15.35) | 285 (81.53) | 0.71 (0.41-1.21) | 0.21 | 0.93 (0.41-2.12) | 0.87 |
| 16th | 245 (10.78) | 224 (92.21) | 1.89 (0.99-3.64) | 0.06 | 1.20 (0.48-2.96) | 0.70 |

[1], Native speaker student considered as missing data because of to low frequence

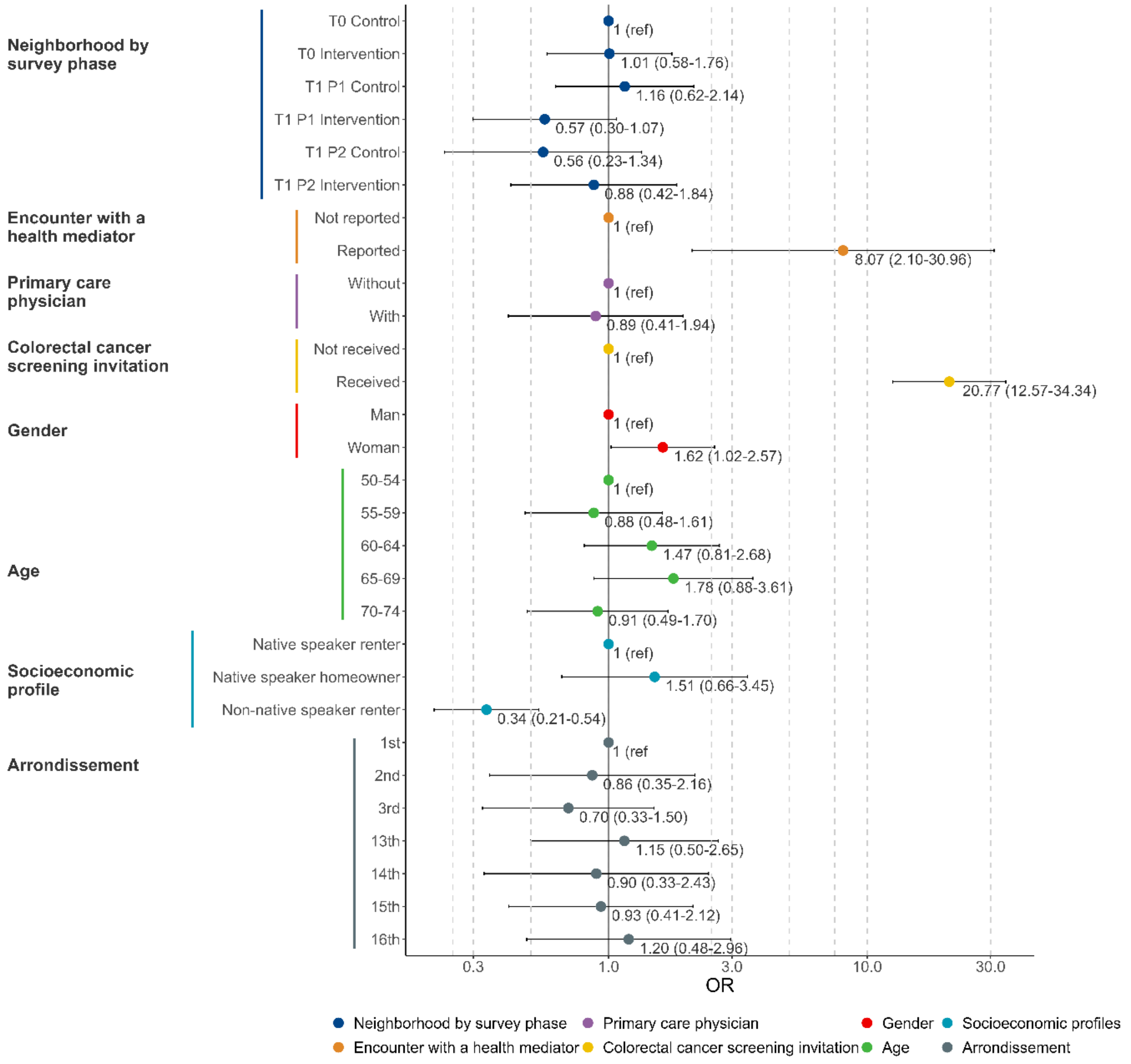

Supplementary figure 16. Factors associated with awareness about colorectal cancer screening in multivariate analysis (n=1959).

#### 9.2. Breast cancer screening awareness

Supplementary table 11. Factors associated with awareness about breast cancer screening.

| | n (%) | n aware (weighted %) | Univariate analysis Odds ratio (95% CI) | p value | Multivariate analysis Odds ratio (95% CI) | p value |
|---|---|---|---|---|---|---|
| **Breast cancer screening awareness** (women aged 50–74 years, n [without missing outcome] = 1432, n [multivariate analysis] =1282) | | | | | | |
| **Neighborhoods by survey phase** | | | | | | |
| T0 Control | 392 (27.37) | 379 (96.60) | 1 (ref) | .. | 1 (ref) | .. |
| T0 Intervention | 335 (23.39) | 312 (93.38) | 0.50 (0.21-1.16) | 0.10 | 0.85 (0.28-2.54) | 0.77 |
| T1 P1 Control | 344 (24.02) | 338 (98.75) | 2.78 (0.92-8.37) | 0.07 | 2.59 (0.81-8.27) | 0.11 |
| T1 P1 Intervention | 188 (13.13) | 177 (94.81) | 0.64 (0.24-1.69) | 0.37 | 0.52 (0.17-1.60) | 0.25 |
| T1 P2 Control | 31 (2.16) | 30 (97.55) | 1.40 (0.17-11.78) | 0.76 | 1.18 (0.12-11.98) | 0.89 |
| T1 P2 Intervention | 130 (9.08) | 120 (91.68) | 0.39 (0.15-1.04) | 0.06 | 0.58 (0.18-1.88) | 0.36 |
| Missing data | 12 (0.84) | 12 (100.00) | .. | .. | .. | .. |
| **Encounter with a health mediator** | | | | | | |
| Not reported | 1333 (93.09) | 1272 (96.55) | 1 (ref) | .. | 1 (ref) | .. |
| Reported | 62 (4.33) | 61 (98.95) | 3.38 (0.45-25.28) | 0.23 | 3.24 (0.43-24.16) | 0.25 |
| Missing data | 37 (2.58) | 35 (96.95) | .. | .. | .. | .. |
| **Primary care physician** | | | | | | |
| Without | 68 (4.75) | 58 (89.28) | 1 (ref) | .. | 1 (ref) | .. |
| With | 1361 (95.04) | 1307 (97.05) | 3.95 (1.64-9.54) | 0.0023 | 2.77 (1.03-7.48) | 0.04 |
| Missing data | 3 (0.21) | 3 (100.00) | .. | .. | .. | .. |
| **Screening invitation** | | | | | | |
| Not received | 193 (13.48) | 175 (91.49) | 1 (ref) | .. | 1 (ref) | .. |
| Received | 1139 (79.54) | 1110 (98.08) | 4.74 (2.17-10.37) | 0.0001 | 4.08 (2.02-8.27) | 0.0001 |
| Missing data | 100 (6.98) | 83 (87.22) | .. | .. | .. | .. |
| **Age** | | | | | | |
| 50-54 | 383 (26.75) | 363 (96.79) | 1 (ref) | .. | 1 (ref) | .. |
| 55-59 | 282 (19.69) | 270 (96.71) | 0.98 (0.35-2.75) | 0.96 | 0.90 (0.29-2.74) | 0.85 |
| 60-64 | 271 (18.92) | 257 (95.86) | 0.77 (0.32-1.84) | 0.55 | 0.52 (0.21-1.30) | 0.16 |
| 65-69 | 229 (15.99) | 224 (97.68) | 1.39 (0.38-5.06) | 0.61 | 0.83 (0.20-3.49) | 0.80 |
| 70-74 | 267 (18.65) | 254 (96.27) | 0.86 (0.38-1.96) | 0.71 | 0.80 (0.23-2.74) | 0.72 |
| **Socioeconomic profile** | | | | | | |
| NS renter | 679 (47.42) | 661 (97.63) | 1 (ref) | .. | 1 (ref) | .. |
| NS homeowner | 206 (14.39) | 202 (98.84) | 2.06 (0.58-7.36) | 0.26 | 1.72 (0.39-7.50) | 0.47 |
| Non-NS renter | 534 (37.29) | 492 (94.05) | 0.38 (0.18-0.82) | 0.01 | 0.39 (0.16-0.96) | 0.04 |
| Missing data[1] | 13 (0.91) | 13 (100.00) | .. | .. | .. | .. |
| **Arrondissement** | | | | | | |
| 1st | 150 (10.47) | 139 (96.06) | 1 (ref) | .. | 1 (ref) | .. |
| 2nd | 182 (12.71) | 175 (94.69) | 0.73 (0.23-2.30) | 0.59 | 0.68 (0.19-2.41) | 0.55 |
| 3rd | 382 (26.68) | 371 (98.45) | 2.62 (0.99-6.88) | 0.05 | 3.37 (1.11-10.27) | 0.03 |
| 13th | 228 (15.92) | 219 (96.86) | 1.27 (0.49-3.28) | 0.63 | 1.99 (0.60-6.62) | 0.26 |
| 14th | 118 (8.24) | 116 (98.71) | 3.14 (0.59-16.69) | 0.18 | 5.66 (0.64-50.43) | 0.12 |
| 15th | 218 (15.22) | 199 (91.09) | 0.42 (0.18-0.97) | 0.04 | 0.82 (0.28-2.40) | 0.71 |
| 16th | 154 (10.75) | 149 (96.51) | 1.14 (0.36-3.58) | 0.83 | 1.02 (0.27-3.80) | 0.98 |

[1], Native speaker student considered as missing data because of to low frequence

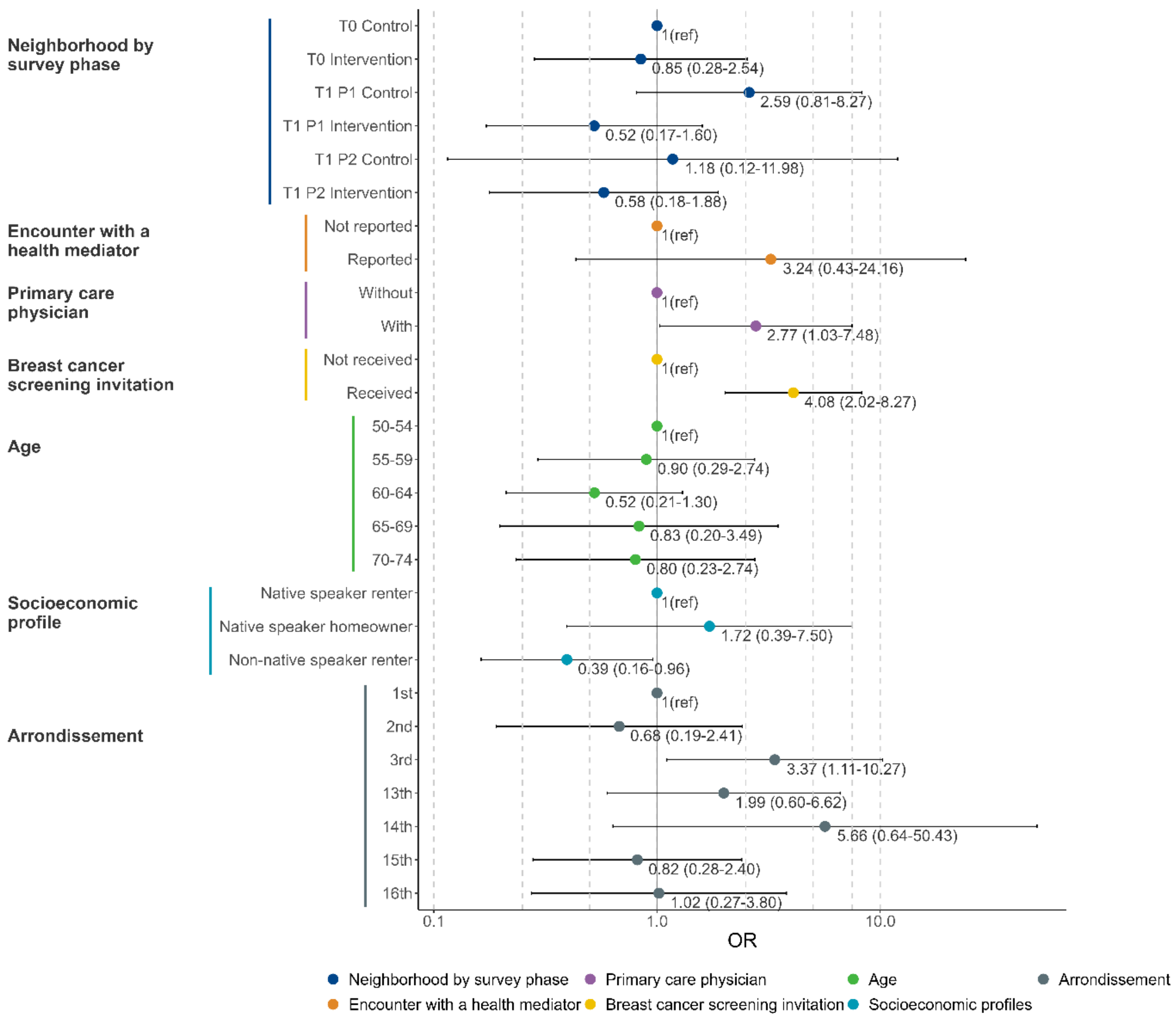

Supplementary figure 17. Factors associated with awareness about breast cancer screening in multivariate analysis (n=1282).

#### 9.3. Cervical cancer screening awareness

Supplementary table 12. Factors associated with awareness about cervical cancer screening.

| | n (%) | n aware (weighted %) | Univariate analysis Odds ratio (95% CI) | p value | Multivariate analysis Odds ratio (95% CI) | p value |
|---|---|---|---|---|---|---|
| **Cervical cancer screening awareness** (women aged 25–64 years, n [without missing outcome] = 2102, n [multivariate analysis] = 1727) | | | | | | |
| **Neighborhoods by survey phase** | | | | | | |
| T0 Control | 618 (29.40) | 524 (83.95) | 1 (ref) | .. | 1 (ref) | .. |
| T0 Intervention | 577 (27.45) | 472 (81.55) | 0.85 (0.59-1.20) | 0.35 | 1.09 (0.69-1.74) | 0.71 |
| T1 P1 Control | 427 (20.31) | 388 (91.49) | 2.06 (1.26-3.34) | 0.0037 | 2.10 (1.21-3.66) | 0.0087 |
| T1 P1 Intervention | 263 (12.51) | 237 (90.36) | 1.79 (1.08-2.97) | 0.02 | 1.64 (0.86-3.11) | 0.13 |
| T1 P2 Control | 39 (1.86) | 32 (81.86) | 0.86 (0.35-2.11) | 0.75 | 0.41 (0.15-1.14) | 0.09 |
| T1 P2 Intervention | 157 (7.47) | 139 (89.31) | 1.60 (0.88-2.89) | 0.12 | 1.31 (0.57-3.04) | 0.53 |
| Missing data | 21 (1.00) | 17 (90.84) | .. | .. | .. | .. |
| **Encounter with a health mediator** | | | | | | |
| Not reported | 1983 (94.34) | 1700 (86.36) | 1 (ref) | .. | 1 (ref) | .. |
| Reported | 88 (4.19) | 82 (94.66) | 2.80 (1.06-7.37) | 0.04 | 2.24 (0.64-7.85) | 0.21 |
| Missing data | 31 (1.47) | 27 (93.50) | .. | .. | .. | .. |
| **Primary care physician** | | | | | | |
| Without | 175 (8.33) | 130 (78.16) | 1 (ref) | .. | 1 (ref) | .. |
| With | 1924 (91.53) | 1677 (87.67) | 1.99 (1.23-3.20) | 0.0047 | 1.19 (0.66-2.16) | 0.56 |
| Missing data | 3 (0.14) | 2 (72.69) | .. | .. | .. | .. |
| **Screening invitation** | | | | | | |
| Not received | 685 (32.59) | 521 (76.54) | 1 (ref) | .. | 1 (ref) | .. |
| Received | 1086 (51.67) | 1031 (94.28) | 5.05 (3.34-7.64) | <0.0001 | 4.30 (2.78-6.65) | <0.0001 |
| Missing data | 331 (15.75) | 257 (81.75) | .. | .. | .. | .. |
| **Age** | | | | | | |
| 25-29 | 274 (13.04) | 226 (83.52) | 1 (ref) | .. | 1 (ref) | .. |
| 30-34 | 234 (11.13) | 195 (85.13) | 1.13 (0.63-2.01) | 0.68 | 1.09 (0.52-2.29) | 0.82 |
| 35-39 | 259 (12.32) | 226 (86.27) | 1.24 (0.65-2.37) | 0.52 | 1.08 (0.49-2.37) | 0.85 |
| 40-44 | 188 (8.94) | 159 (83.09) | 0.97 (0.51-1.85) | 0.93 | 0.86 (0.38-1.97) | 0.73 |
| 45-49 | 213 (10.13) | 185 (88.83) | 1.57 (0.78-3.15) | 0.20 | 1.79 (0.72-4.47) | 0.21 |
| 50-54 | 379 (18.03) | 336 (89.10) | 1.61 (0.90-2.90) | 0.11 | 1.72 (0.80-3.69) | 0.16 |
| 55-59 | 283 (13.46) | 249 (91.21) | 2.05 (1.14-3.66) | 0.02 | 2.32 (1.00-5.39) | 0.05 |
| 60-64 | 272 (12.94) | 233 (85.85) | 1.20 (0.66-2.17) | 0.55 | 1.18 (0.54-2.59) | 0.67 |
| **Socioeconomic profile** | | | | | | |
| NS renter | 947 (45.05) | 870 (92.59) | 1 (ref) | .. | 1 (ref) | .. |
| NS homeowner | 197 (9.37) | 188 (95.62) | 1.75 (0.72-4.22) | 0.22 | 2.34 (0.88-6.19) | 0.09 |
| NS student | 105 (5.00) | 86 (78.99) | 0.30 (0.15-0.60) | 0.0007 | 0.61 (0.25-1.47) | 0.27 |
| Non-NS renter | 852 (40.53) | 664 (77.63) | 0.28 (0.19-0.41) | <0.0001 | 0.26 (0.16-0.44) | <0.0001 |
| Missing data | 1 (0.05) | 1 (100.00) | .. | .. | .. | .. |
| **Arrondissement** | | | | | | |
| 1st | 225 (10.70) | 192 (86.62) | 1 (ref) | .. | 1 (ref) | .. |
| 2nd | 236 (11.23) | 217 (92.37) | 1.87 (0.85-4.12) | 0.12 | 2.07 (0.85-5.05) | 0.11 |
| 3rd | 572 (27.21) | 472 (84.74) | 0.86 (0.49-1.51) | 0.60 | 1.12 (0.55-2.28) | 0.75 |
| 13th | 333 (15.84) | 288 (86.44) | 0.98 (0.54-1.80) | 0.96 | 1.39 (0.65-2.97) | 0.40 |
| 14th | 188 (8.94) | 164 (82.67) | 0.74 (0.37-1.47) | 0.39 | 0.66 (0.31-1.44) | 0.30 |
| 15th | 315 (14.99) | 258 (83.69) | 0.79 (0.44-1.43) | 0.44 | 1.40 (0.64-3.07) | 0.39 |
| 16th | 233 (11.08) | 218 (94.23) | 2.52 (1.20-5.31) | 0.01 | 2.90 (1.17-7.19) | 0.02 |

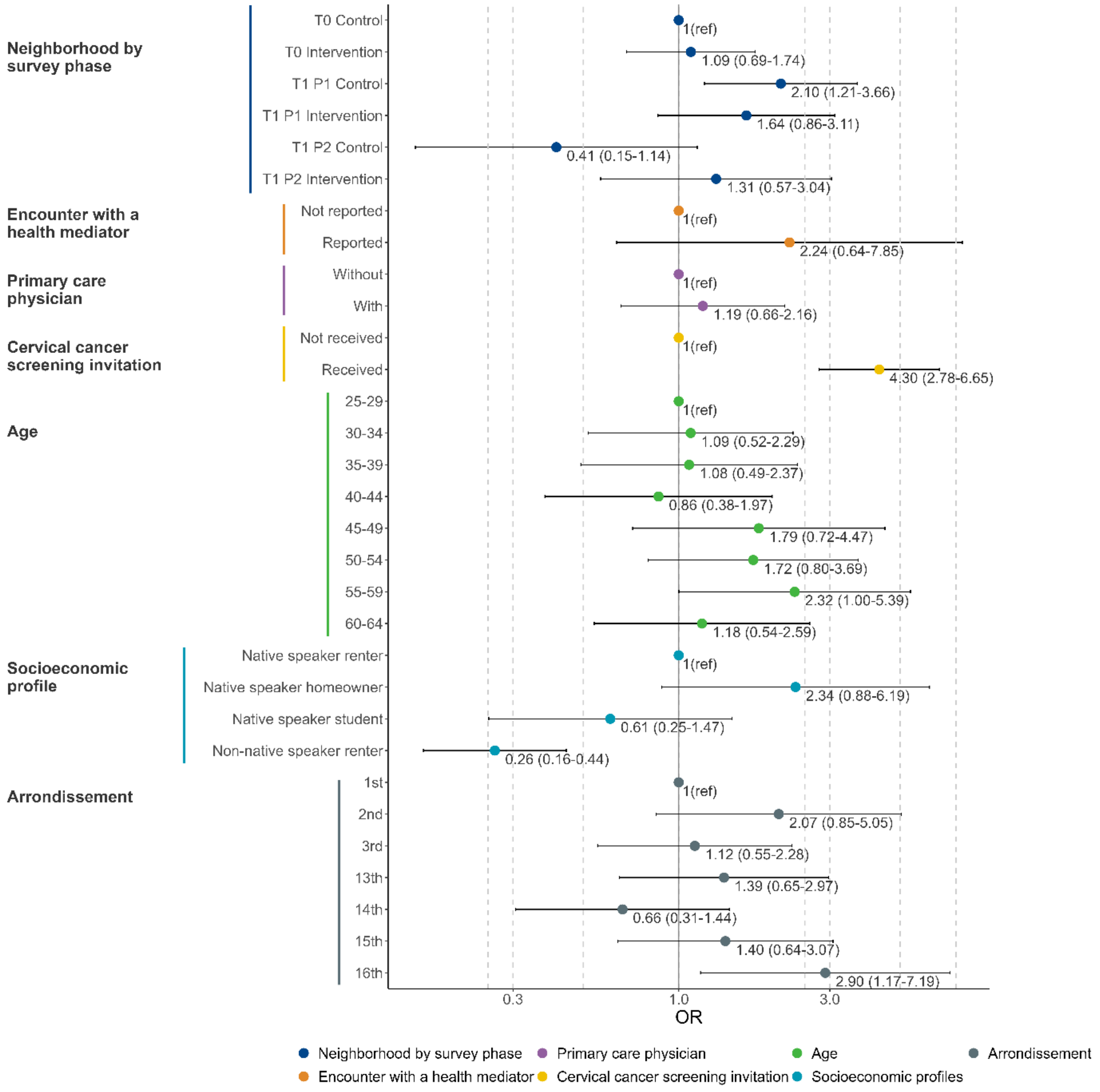

Supplementary figure 18. Factors associated with awareness about cervical cancer screening in multivariate analysis (n=1727).
